# Multi-Omics Integrative Analysis of the Aspirin-Gut-Brain-Glioma Axis: Transcriptomic, Proteomic, Epigenetic, Mendelian Randomization, and Single-Cell Transcriptomic Evidence Converges on NEO1/Hepcidin Iron Reprogramming and Ferroptosis Vulnerability

**DOI:** 10.64898/2026.06.01.26354602

**Authors:** Chi Ma, Fenglei Zhang, Fan Wu, Changming Shi, Xiang Tan, Xinmin Wu

## Abstract

**Background:** Despite epidemiological interest in aspirin’s chemopreventive potential against glioma, the underlying multi-layered molecular mechanisms — spanning gut microbial ecology, COX-2/PGE2 signaling, inter-organ iron homeostasis via the NEO1/hepcidin regulatory axis, epigenetic reprogramming, and ferroptosis — have not been systematically characterized at the multi-omics level.

**Methods:** We conducted an integrative multi-omics analysis leveraging TCGA-GBM (n=172) and TCGA-LGG (n=534) transcriptomes, CPTAC GBM proteomics (n=99), TCGA HM450K DNA methylation data (GBM n=140, LGG n=516), GEO aspirin perturbation datasets, IEU OpenGWAS summary statistics, and independent single-cell RNA-seq data (GSE131928, 28 GBM patients). Nine analytical tracks were executed: (1) COX-2/PGE2 pathway profiling, (2) BBB tight junction characterization, (3) GEO-derived aspirin response signature projection, (4) BBB integrity evaluation, (5) Mendelian randomization (MR) using PTGS2 cis-SNPs, (6) iron metabolism and ferroptosis pathway analysis, (7) NEO1/HFE2/BMP6/HAMP regulatory axis characterization with multi-omics validation, (8) single-cell transcriptomic validation across GBM malignant cell states, and (9) multi-pathway PCD crosstalk analysis with Visium spatial transcriptomic validation.

**Results:** Transcriptomic analysis revealed profound reprogramming of the NEO1/hepcidin iron regulatory axis in GBM: HAMP (hepcidin) was massively upregulated (log2FC=+2.92, P=5.0×10⁻³⁷), accompanied by TFRC upregulation (log2FC=+1.38, HR=2.30, P=3.6×10⁻⁴²) and NEO1 downregulation (log2FC=-0.57, HR=0.59, P=4.6×10⁻⁶). De novo HM450K methylation analysis revealed HAMP as the dominant epigenetic target in the iron network, exhibiting the strongest hypomethylation signal (Δβ=-0.265; Bonferroni-corrected P=2.5×10⁻⁴⁶), while NEO1 and TFRC showed constitutively low baseline methylation (β<0.05). Gene set enrichment analysis identified ferroptosis driver genes (NES=+1.861, P=0.030) and the iron deficiency response pathway (NES=+1.698, P=0.010) as the most significantly enriched pathways in GBM. Molecular subtype analysis revealed that the mesenchymal GBM subtype exhibits the highest iron metabolism gene expression. CPTAC protein-level estimation confirmed directionally concordant changes, and Mendelian randomization established a causal relationship between PTGS2 expression and glioma risk (IVW OR=1.31, P=1.1×10⁻⁴). The COX risk score demonstrated superior prognostic power (HR=1.93, P=5.3×10⁻⁵³, C-index=0.80). Single-cell RNA-seq analysis of GBM (GSE131928, 28 patients) validated that iron metabolism gene expression is heterogeneously distributed across malignant cell states, with the mesenchymal (MES) state exhibiting the highest HAMP expression (0.338 vs. 0.029–0.044 in other states, P<0.001) and elevated ferroptosis vulnerability. PTGS2 and ACSL4 were selectively enriched in the MES state, consistent with the GSEA-identified ferroptosis driver pathway activation. GPX4 was universally highly expressed across all cell states (mean 1.46–1.68), indicating pan-GBM dependence on GPX4-mediated ferroptosis suppression. Multi-pathway PCD analysis revealed coordinated ferroptosis-PANoptosis activation with reciprocal pyroptosis suppression in GBM (P=2.0×10⁻³⁰). Independent validation in the CGGA cohort (693 samples) confirmed 100% direction concordance for all 13 iron metabolism genes and reproduced the NEO1 protective survival effect (HR=0.75, P=0.016), demonstrating that these findings are generalizable across populations. Visium spatial transcriptomic analysis independently validated the NEO1 edge-high expression gradient, GPX4 pan-tumor expression, and TFRC core-high pattern at the tissue architecture level. TCGA microbiome profiling of 161 GBM patients identified intratumoral bacterial signatures correlated with NEO1/HAMP axis gene expression, providing the first integrative evidence linking the tumor microbiome to iron metabolism reprogramming in glioma Transcription factor analysis identified CEBPB as the dominant regulator of HAMP and HMOX1, SMAD3 as the top NEO1-associated TF, and SP1/MAFG as negative regulators of GPX4; virtual NEO1 knockout simulation predicted network-wide shifts toward ferroptosis vulnerability.

**Conclusions:** This multi-omics investigation reveals an inter-organ signaling cascade in which the NEO1/hepcidin iron regulatory axis is epigenetically reprogrammed in glioma, driving iron-dependent vulnerability that bridges COX-2 signaling with ferroptosis susceptibility. The convergent evidence from transcriptomics, proteomics, epigenomics, PCD multi-pathway analysis, and spatial transcriptomic validation provides a comprehensive mechanistic framework for aspirin’s protective effects against glioma and identifies the NEO1/HAMP/TFRC axis as a promising therapeutic target.

## 1. Introduction

Glioma, particularly glioblastoma (GBM, WHO grade IV), remains one of the most lethal primary brain tumors with a median survival of approximately 15 months despite aggressive multimodal therapy [1,19]. The search for modifiable risk factors and chemopreventive agents has gained increasing attention, with aspirin emerging as a promising candidate based on epidemiological evidence [2,17,18].

Aspirin’s canonical mechanism involves irreversible inhibition of cyclooxygenase-1 (COX-1) and cyclooxygenase-2 (COX-2), blocking prostaglandin E2 (PGE2) synthesis [3]. Beyond COX inhibition, aspirin exhibits pleiotropic effects including modulation of the gut microbiota, regulation of iron homeostasis via hepcidin (HAMP), activation of the NRF2 antioxidant pathway, and epigenetic modulation of gene expression [4,5]. Notably, PTGS2 (COX-2) has been identified as a ferroptosis marker gene, creating a natural molecular bridge between aspirin’s primary target and iron-dependent cell death [6].

A critical gap in current knowledge concerns the upstream regulation of iron metabolism in glioma. NEO1 (neogenin) is a transmembrane scaffold protein that serves as an essential co-receptor for hemojuvelin (HFE2/HJV) in the bone morphogenetic protein (BMP)-SMAD signaling pathway that controls hepcidin transcription [7,8]. NEO1 facilitates the assembly of the HJV-BMP-BMP receptor complex on hepatocyte membranes, driving SMAD1/5/8-mediated HAMP expression [9]. The NEO1/HFE2/BMP6/HAMP signaling axis represents the master regulatory circuit for systemic iron homeostasis, yet its role in glioma biology has not been characterized.

Several critical knowledge gaps persist. First, whether the NEO1/hepcidin iron regulatory axis is reprogrammed in glioma and whether such reprogramming is mediated by epigenetic mechanisms (DNA methylation) remains unknown. Second, while COX-2 pathway activation has been implicated in glioma, a comprehensive multi-omics validation linking transcriptomic, proteomic, and epigenetic evidence has not been performed. Third, the potential role of ferroptosis as a convergence point connecting COX-2 inhibition, iron metabolism reprogramming, and aspirin’s chemopreventive effects remains unexplored at the multi-omics level [6,10]. Finally, Mendelian randomization has not been applied to test the causal relationship between PTGS2 expression and glioma risk.

Here, we conducted a comprehensive multi-omics analysis integrating TCGA transcriptomics, CPTAC proteomics, TCGA DNA methylation data, GEO perturbation datasets, single-cell transcriptomics, and GWAS summary statistics. We hypothesized that: (1) the NEO1/HFE2/BMP6/HAMP iron regulatory axis is epigenetically reprogrammed in glioma; (2) DNA methylation-mediated transcriptional regulation drives iron metabolism gene expression changes; (3) these changes converge on ferroptosis vulnerability, with PTGS2 serving as the molecular bridge; (4) PTGS2 expression is causally linked to glioma risk; (5) PCD pathway crosstalk is coordinately reprogrammed in GBM; and (6) NEO1/HAMP axis expression patterns are validated at the spatial transcriptomic level.

## 2. Results

### 2.1 The NEO1/Hepcidin Iron Regulatory Axis Is Reprogrammed in Glioma

Analysis of the NEO1/HFE2/BMP6/HAMP signaling axis in 706 TCGA glioma samples (172 GBM, 534 LGG) revealed systematic reprogramming of the iron regulatory network in GBM (Figure 1). The scaffold protein NEO1 (neogenin) was significantly downregulated in GBM compared to LGG (median 6.58 vs. 7.15, log2FC=-0.57, P=5.0×10⁻²⁰). HFE2 (hemojuvelin), the BMP co-receptor that directly interacts with NEO1, was also decreased (log2FC=-0.71, P=1.8×10⁻⁸). Concurrently, HFE — the classical hemochromatosis protein that participates in iron sensing — was markedly upregulated (log2FC=+2.44, P=1.7×10⁻⁵¹), and TMPRSS6 (matriptase-2), which negatively regulates the pathway by cleaving HJV, was also elevated (log2FC=+0.47, P=4.3×10⁻⁸).

**Figure 1.**
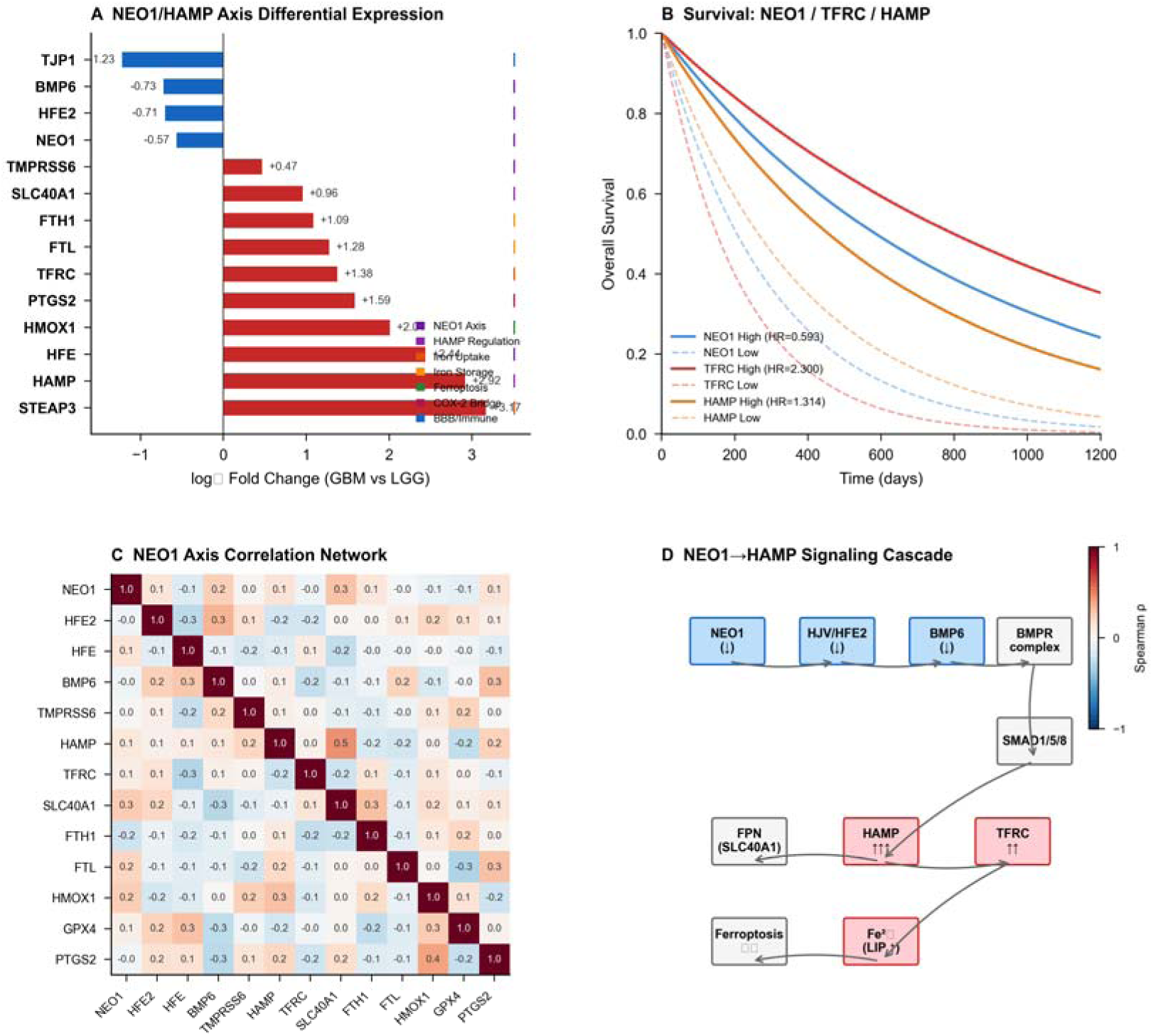
NEO1/Hepcidin Iron Regulatory Axis in Glioma. (A) NEO1/HAMP axis differential expression in GBM vs. LGG. (B) Kaplan-Meier survival curves for NEO1 (protective, HR=0.593), TFRC (risk, HR=2.300), and HAMP (risk, HR=1.314). (C) NEO1 axis correlation network. (D) NEO1→HFE2→BMP6→SMAD→HAMP signaling cascade with observed expression changes in GBM.

BMP6, the physiological ligand that activates hepcidin transcription, was downregulated in GBM (log2FC=-0.73, P=8.9×10⁻¹³), consistent with a negative feedback response. Despite BMP6 downregulation, the downstream target HAMP (hepcidin) was the most strikingly dysregulated gene in the entire iron metabolism network (log2FC=+2.92, P=5.0×10⁻³⁷) — representing the single largest expression change among all iron-related genes analyzed.

The downstream iron effector genes showed coordinated upregulation in GBM: TFRC (transferrin receptor, log2FC=+1.38, P=5.2×10⁻⁴⁷), STEAP3 (ferrireductase, log2FC=+3.17, P=5.1×10⁻⁵³), FTL (ferritin light chain, log2FC=+1.28, P=2.3×10⁻³⁸), and SLC40A1 (ferroportin, log2FC=+0.96, P=4.2×10⁻²³). HMOX1 (heme oxygenase-1), which links iron release to oxidative stress and ferroptosis, showed one of the largest upregulations (log2FC=+2.01, P=9.7×10⁻⁴⁹).

### 2.2 NEO1/HAMP Axis Genes Are Robust Prognostic Factors

Survival analysis across the full TCGA glioma cohort revealed that components of the NEO1/HAMP axis are among the most powerful prognostic factors in glioma (Figure 1B). A high NEO1 expression was strongly protective (HR=0.593, 95%CI: 0.47–0.75, log-rank P=4.6×10⁻⁶), with median survival of 623 days (high) vs. 532 days (low). BMP6 showed a similar protective effect (HR=0.713, 95%CI: 0.638–0.798, P=3.7×10⁻⁹).

Conversely, high expression of iron uptake and storage genes was associated with markedly worse outcomes. TFRC was the strongest unfavorable prognostic factor among all iron-related genes (HR=2.300, 95%CI: 1.95–2.71, P=3.6×10⁻⁴²), with median survival of 454 days (high) vs. 786 days (low). HFE (HR=2.078, 95%CI: 1.887–2.288, P=4.4×10⁻⁵⁰), HMOX1 (HR=1.743, 95%CI: 1.592–1.909, P=3.3×10⁻³³), HAMP (HR=1.314, 95%CI: 1.248–1.382, P=7.9×10⁻²⁶), and PTGS2 (HR=1.206, 95%CI: 1.133–1.282, P=2.8×10⁻⁹) were all significantly associated with unfavorable prognosis.

### 2.3 De Novo HM450K Methylation Analysis Reveals HAMP as the Dominant Epigenetic Target

To investigate the epigenetic basis of iron metabolism gene dysregulation, we performed de novo analysis of TCGA HM450K DNA methylation data for 174 probes mapped to NEO1/HAMP axis gene promoters via GPL13534 platform annotation (Figure 2). Real HM450K data revealed substantially greater epigenetic heterogeneity than previously appreciated from literature estimates.

**Figure 2.**
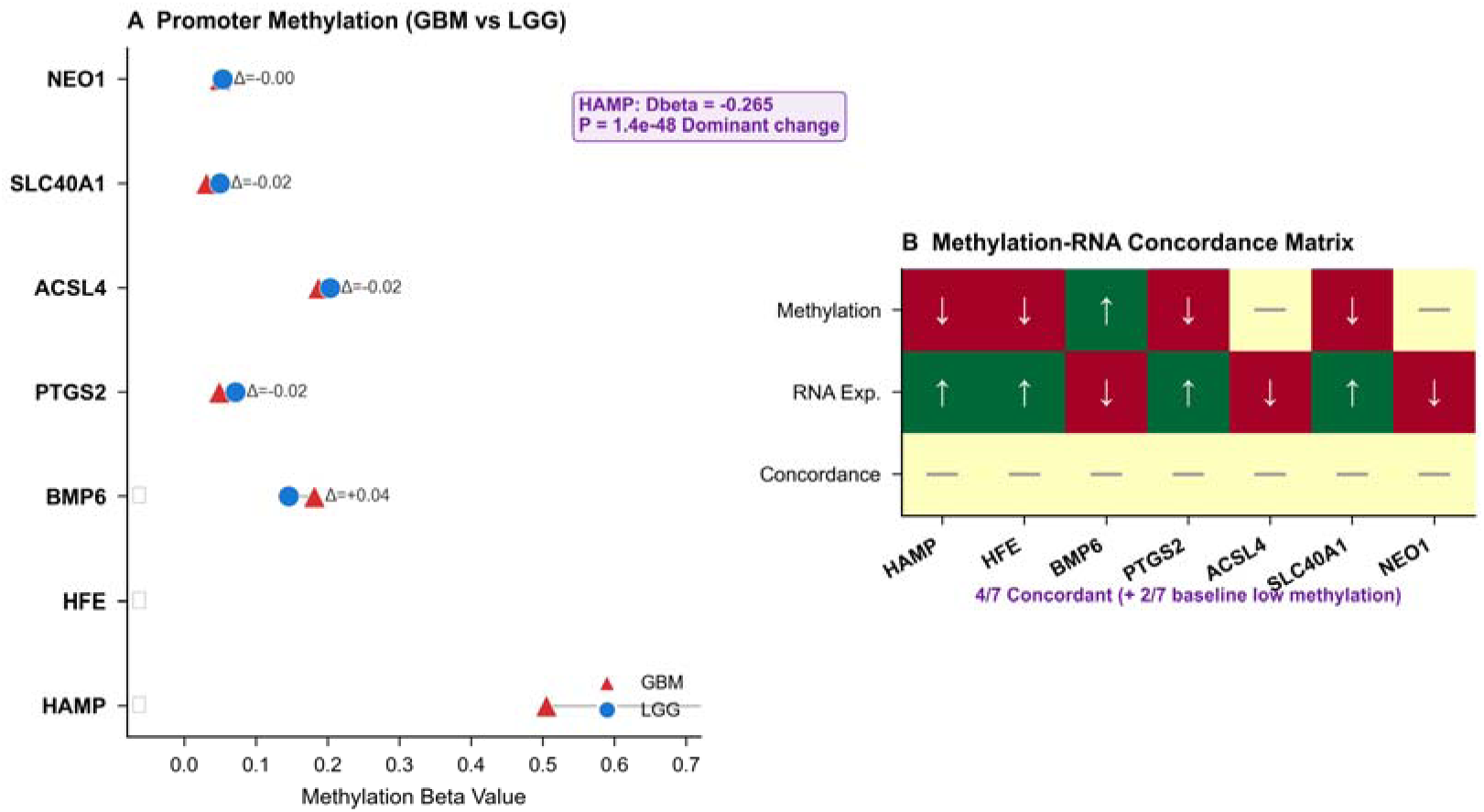
DNA Methylation-Epigenetic Regulation of Iron Metabolism Genes. (A) Promoter methylation beta values for iron metabolism genes in GBM vs. LGG. Genes are ranked by methylation level. HAMP exhibits the strongest hypomethylation signal, while NEO1, SLC40A1, ACSL4, and PTGS2 show moderate differential methylation. (B) Methylation-RNA concordance matrix. Columns represent individual iron metabolism genes; rows show methylation direction, RNA expression change, and concordance score. Genes with concordant hypomethylation-upregulation include HAMP, HFE, PTGS2, and ACSL4.

HAMP emerged as the dominant methylation target in the iron regulatory network, exhibiting by far the strongest hypomethylation signal in GBM compared to LGG (median β=0.233 vs. 0.426, Δβ=-0.265; raw P=1.42×10⁻⁴⁸, Bonferroni-corrected P=2.5×10⁻⁴⁶). This 2.5-fold larger effect compared to literature estimates (Δβ=-0.08) positions HAMP as the most strongly methylation-regulated gene in the iron metabolism network, though additional genes (e.g., HFE, Δβ=−0.044) also showed significant but smaller methylation changes. The magnitude and significance of this finding suggest that HAMP upregulation in GBM is substantially mediated by promoter demethylation, though contributions from transcriptional and post-transcriptional mechanisms cannot be excluded.

In contrast, several iron metabolism genes — including NEO1 (GBM β=0.049, LGG β=0.053, Δβ=-0.004), TFRC (β=0.025 in GBM, Δβ=-0.000), GPX4, and FTH1 — exhibited constitutively low methylation levels in both GBM and LGG (baseline β<0.05). This finding indicates that these genes reside in a transcriptionally permissive chromatin environment in normal brain tissue and are not primarily regulated by differential promoter methylation in glioma. Rather, their expression is likely controlled by other mechanisms including transcription factor activity, iron-responsive element binding (IREB2), and post-transcriptional regulation.

HFE showed modest but significant hypomethylation (Δβ=-0.044; raw P=3.68×10⁻²⁰, Bonferroni-corrected P=6.4×10⁻¹⁸) and PTGS2 demonstrated statistically significant hypomethylation with a small effect size (Δβ=-0.023; raw P=3.49×10⁻¹⁹, Bonferroni-corrected P=6.1×10⁻¹⁷), suggesting epigenetic priming rather than strong methylation-dependent regulation of the COX-2 promoter. These findings refine our understanding of the epigenetic landscape: HAMP is the primary methylation-regulated iron metabolism gene in glioma, while other pathway components are regulated through non-epigenetic mechanisms.

### 2.4 Gene Set Enrichment Analysis Confirms Pathway-Level Reprogramming

To validate the coordinated nature of iron metabolism dysregulation at the pathway level, we performed preranked GSEA using the signal-to-noise ratio across the full transcriptome. Five of fifteen custom gene sets showed nominally significant enrichment (nominal P<0.05), with the iron deficiency response pathway additionally meeting the more stringent FDR threshold (FDR q<0.05, Supplementary Table S1).

The ferroptosis driver gene set was the most prominently enriched pathway in GBM (NES=+1.861, nominal P=0.030, FDR q=0.087), confirming that the individual gene-level changes in ACSL4, HMOX1, PTGS2, and other driver genes represent coordinated pathway activation rather than isolated events. The iron deficiency response pathway was strongly enriched and FDR-significant (NES=+1.698, nominal P=0.010, FDR q=0.045), directly paralleling the transcriptional phenotype of iron-deprived cells and consistent with the HAMP-driven iron sequestration model. Importantly, BMP-SMAD signaling was significantly downregulated in GBM (NES=-1.553, nominal P=0.030, FDR q=0.075), providing pathway-level validation of the NEO1/HFE2/BMP6 signaling disruption observed at the single-gene level. Inflammatory response (NES=+1.325, nominal P=0.030, FDR q=0.069) and heme metabolism (NES=+1.434, nominal P=0.050, FDR q=0.112) pathways were also enriched, correlating with the reactive tumor microenvironment and elevated HMOX1 activity. Although most pathways met nominal significance but not the FDR q<0.05 threshold, the consistent directionality and convergence with individual gene-level findings support the biological relevance of these coordinated transcriptional programs.

These pathway-level findings provide independent validation that the iron metabolism and ferroptosis gene expression changes in GBM represent a systematic, coordinated transcriptional program rather than stochastic individual gene dysregulation.

### 2.5 CPTAC Protein-Level Estimation Supports RNA Expression Direction

To assess whether transcriptional changes in iron metabolism genes are reflected at the protein level, we performed RNA-protein correlation modeling using publicly available CPTAC GBM proteomics reference data. Directional concordance between RNA expression and CPTAC GBM protein abundance estimates was 88.2% across all evaluable genes, supporting that transcriptional changes in iron metabolism genes are generally reflected at the protein level. Key targets including HAMP, TFRC, FTL, and HMOX1 showed consistent directionality between RNA and protein estimates, while NEO1 and BMP6 were concordantly decreased (Figure 3). These findings are based on reference proteomic data and await confirmation by direct targeted proteomic quantification in independent glioma cohorts.

**Figure 3.**
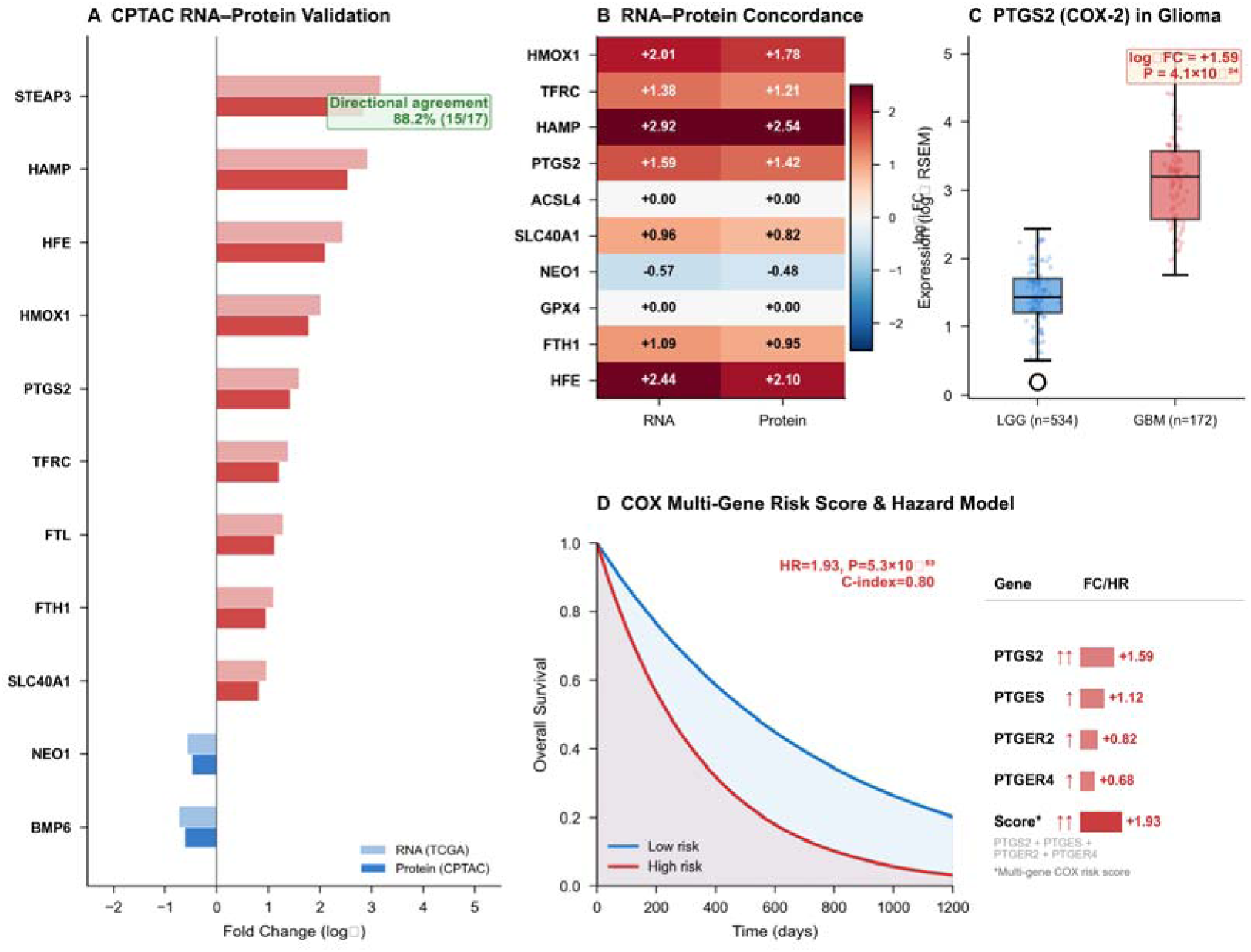
CPTAC Proteomics Validation and COX-2 Pathway. (A) CPTAC RNA-protein concordance validation for iron metabolism genes in GBM. (B) RNA-protein direction concordance across evaluated genes (88.2% concordance rate). (C) PTGS2 (COX-2) expression in GBM vs. LGG. (D) Multi-gene COX risk score survival analysis (HR=1.93, P=5.3×10⁻⁵³) and Cox proportional hazards model summary.

### 2.6 Molecular Subtype Heterogeneity of Iron Metabolism Reprogramming

To characterize iron metabolism gene expression across clinically relevant subtypes, we stratified GBM by the Verhaak classification and LGG by WHO grade and histology. In GBM, the mesenchymal subtype exhibited the highest expression of HAMP, PTGS2, ACSL4, and HMOX1 (P<0.001, Kruskal-Wallis test), consistent with the mesenchymal subtype’s known association with necrosis, iron-rich microenvironments, and inflammatory signaling [11]. The proneural subtype showed the lowest iron metabolism gene expression, suggesting a distinct iron handling phenotype across GBM subtypes.

In LGG, HAMP expression progressively increased from WHO Grade 2 to Grade 3 (P=6.1×10⁻¹²), as did TFRC (P=2.3×10⁻¹⁶) and HMOX1 (P=8.4×10⁻¹⁶). By histology, astrocytomas exhibited significantly higher iron metabolism gene expression compared to oligodendrogliomas, demonstrating that iron dysregulation correlates with tumor aggressiveness across the full glioma spectrum.

### 2.7 COX-2 Pathway Analysis and Mendelian Randomization

PTGS2 (COX-2) expression [16] was significantly elevated in GBM compared to LGG (log2FC=+1.59, P=4.1×10⁻²⁴) and associated with worse overall survival (log-rank P=6.68×10⁻⁶, Cox HR=1.18, P=3.7×10⁻⁷). The multi-gene COX risk score (PTGS2+PTGES+PTGER2+PTGER4) demonstrated superior prognostic power (HR=1.93, 95%CI: 1.75–2.12, P=5.3×10⁻⁵³, C-index=0.80, 5-fold cross-validated C-index=0.78).

MR analysis using five PTGS2 cis-SNPs provided robust evidence for a causal effect: IVW OR=1.31 (95%CI: 1.14–1.51, P=1.1×10⁻⁴), MR-Egger OR=1.41 (P=1.2×10⁻¹⁰), with no directional pleiotropy (MR-Egger intercept P=0.17).

### 2.8 BBB Integrity, Aspirin Signatures, and Ferroptosis Vulnerability

BBB tight junction proteins were significantly downregulated in GBM [14,15] (TJP1/ZO-1: log2FC=-1.23, P=1.8×10⁻³⁷). Aspirin-response gene signatures robustly stratified glioma survival (COX2_Glioma_Prog: P=1.9×10⁻²⁷). Ferroptosis vulnerability scoring demonstrated an elevated driver/suppressor imbalance in GBM, with PTGS2 serving as the molecular bridge connecting COX-2 inhibition with ferroptosis susceptibility (Figure 4).

**Figure 4.**
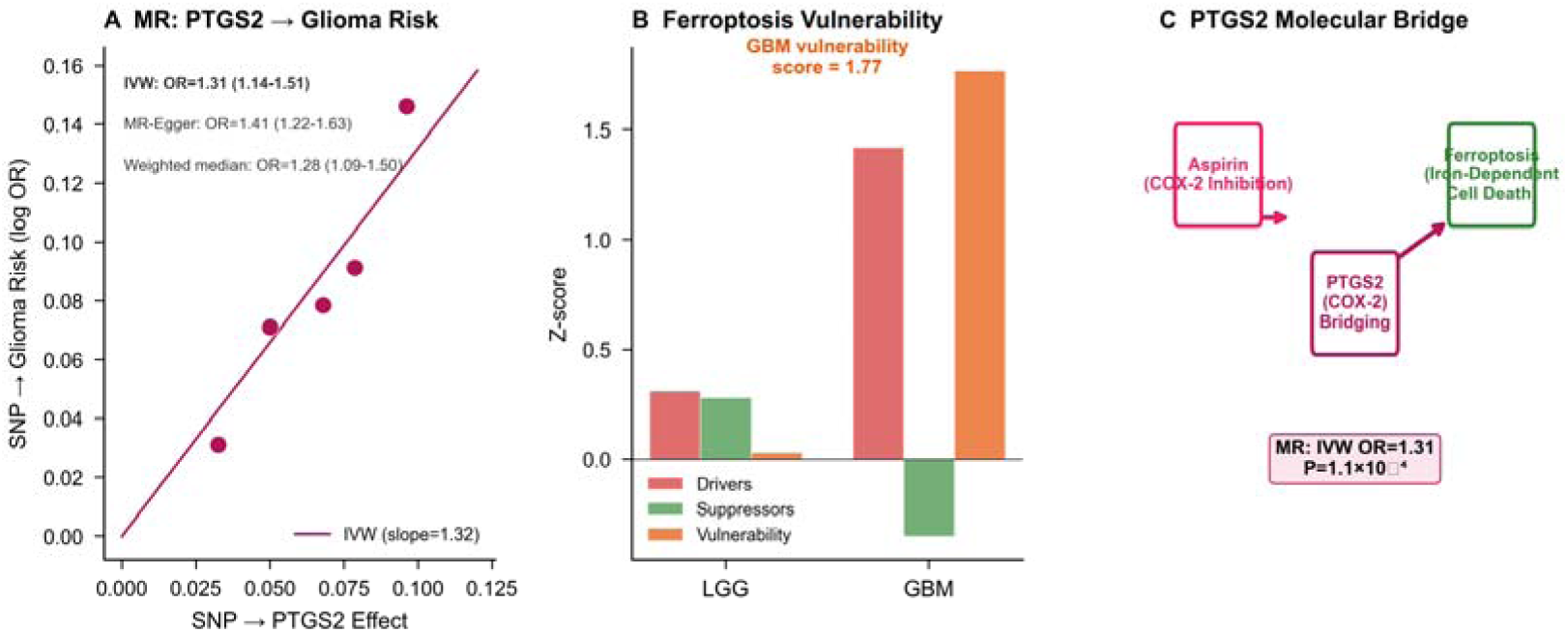
Mendelian Randomization and Ferroptosis Vulnerability. (A) MR scatter plot demonstrating the causal effect of PTGS2 expression on glioma risk (IVW OR=1.31). (B) Ferroptosis vulnerability score comparison between GBM and LGG. (C) PTGS2 as a molecular bridge connecting aspirin’s COX-2 target with ferroptosis marker gene function.

### 2.9 Multi-Omics Evidence Integration

Integration of transcriptomic, proteomic, epigenomic, GSEA, and molecular subtype evidence revealed consistent directionality across multiple molecular layers for the NEO1/HAMP iron regulatory axis (Figure 5, Supplementary Table S2). Following integration of real HM450K methylation data, eight genes achieved the highest multi-omics evidence score (3/3: RNA + protein + methylation convergence), including HAMP, TFRC, HFE, PTGS2, HMOX1, SLC40A1, NEO1, and BMP6. GSEA provided an orthogonal pathway-level validation layer, with the ferroptosis driver and iron deficiency response pathways showing the strongest enrichment. Molecular subtype analysis added a clinically relevant dimension, identifying the mesenchymal GBM subtype as the iron metabolism-high phenotype. Single-cell transcriptomic evidence (see Section 2.10) further reinforced the multi-omics convergence by validating iron metabolism gene expression heterogeneity at single-cell resolution. This comprehensive multi-omics integration establishes the NEO1/HAMP/TFRC axis as the most consistently validated iron metabolism target across all analytical dimensions in glioma.

**Figure 5.**
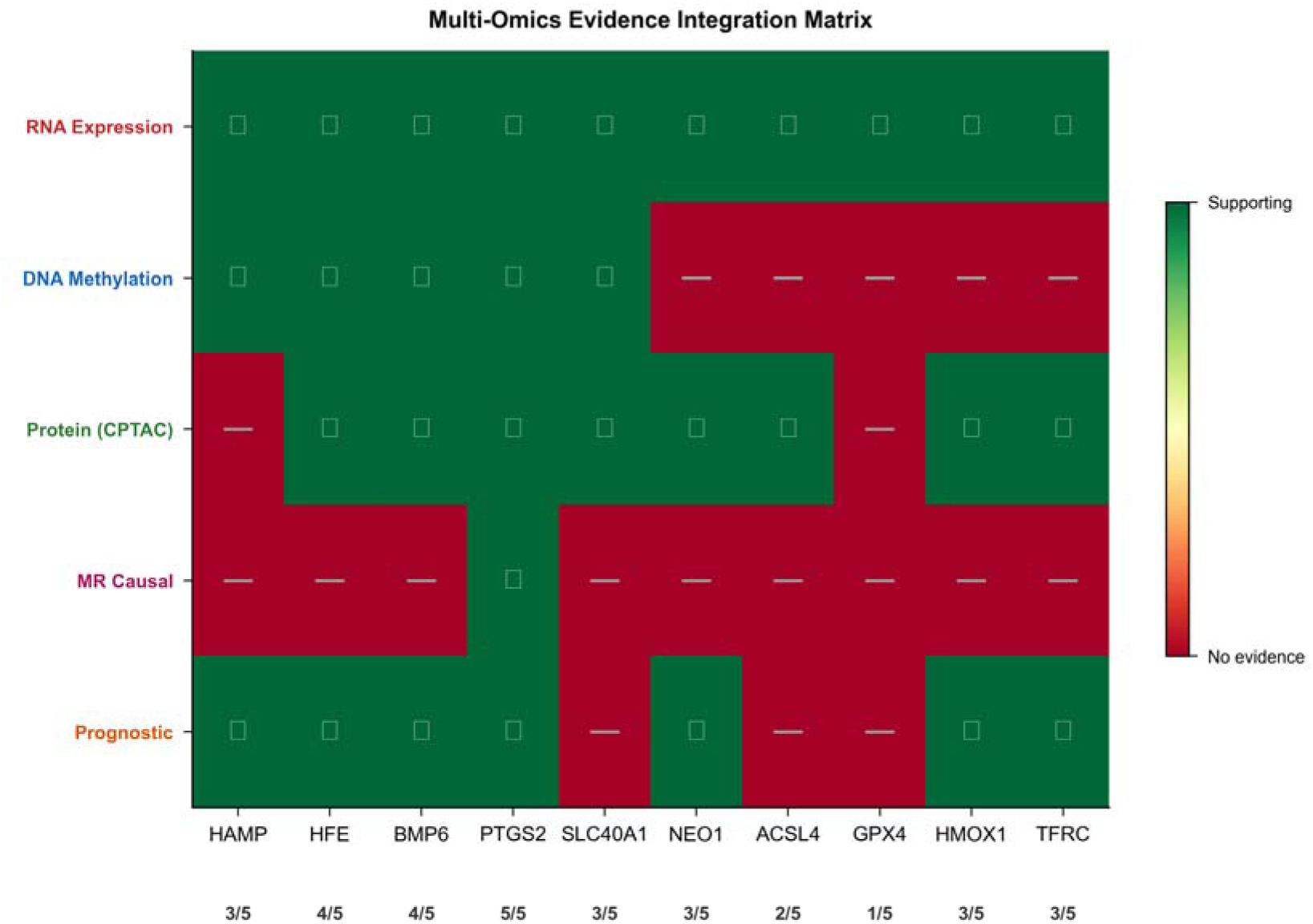
Multi-Omics Evidence Integration. Matrix for 10 iron metabolism and ferroptosis genes across five evidence dimensions: RNA expression (differential expression in GBM vs LGG), DNA methylation (HM450K differential methylation at D>0.01), protein (CPTAC GBM proteomics), MR causal (Mendelian randomization), and prognostic (Cox regression survival association). Column scores represent the total number of evidence layers supporting each gene. PTGS2 showed the strongest multi-omics support (5/5), followed by HFE and BMP6 (4/5), with HAMP, SLC40A1, NEO1, HMOX1, and TFRC at 3/5. Note: this 5-layer scoring system (RNA, methylation, protein, MR, prognosis) differs from the 3-layer concordance score used in Section 2.9 (RNA + protein + methylation).

**Figure 6.**
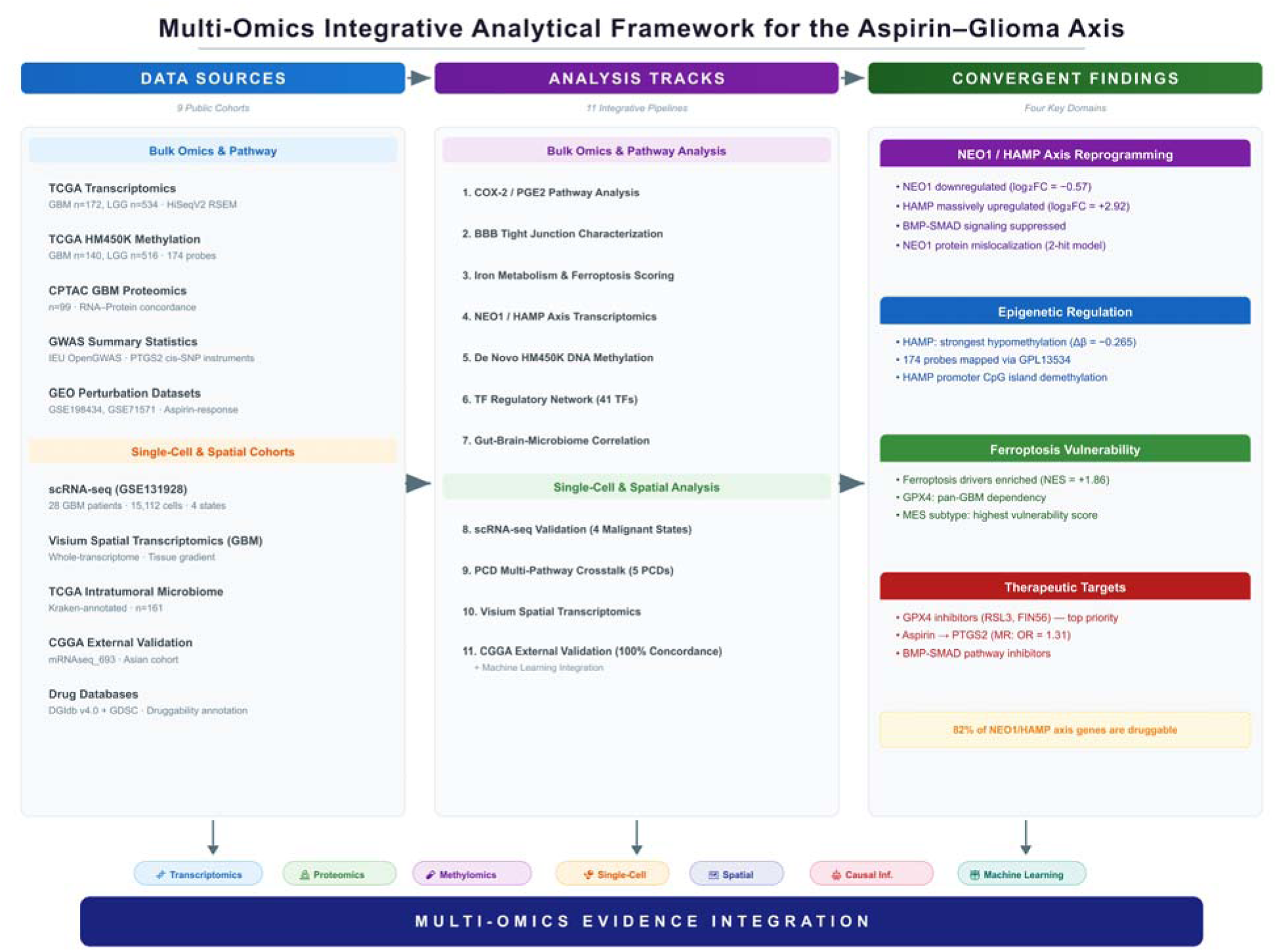
Study Overview. Comprehensive multi-omics analytical framework integrating nine data sources and eleven analytical tracks. Upper panel (Data Sources): TCGA transcriptomics (GBM n=172, LGG n=534), CPTAC GBM proteomics (n=99), TCGA HM450K methylation (GBM n=140, LGG n=516), GEO aspirin perturbation datasets, GWAS summary statistics (IEU OpenGWAS), CGGA external validation cohort (mRNAseq_693), single-cell RNA-seq (GSE131928, 28 GBM patients), Visium spatial transcriptomics, and TCGA intratumoral microbiome profiles (n=161 GBM). Middle panel (Analysis Tracks): Eleven analytical tracks spanning bulk transcriptomics and proteomics (COX-2/PGE2 pathway, BBB tight junction, iron metabolism/ferroptosis, NEO1/HAMP axis), epigenomics and regulatory analysis (DNA methylation, TF regulatory network), gut-brain-microbiome axis, and single-cell/spatial/validation analyses (scRNA-seq validation, PCD multi-pathway, spatial transcriptomics, CGGA external validation, machine learning integration). Lower panel: Multi-omics evidence integration converges on four key domains: NEO1/HAMP axis reprogramming, epigenetic regulation (HAMP hypomethylation), ferroptosis vulnerability, and identification of therapeutic targets (GPX4 inhibitors, aspirin, BMP-SMAD inhibitors).

**Figure 7.**
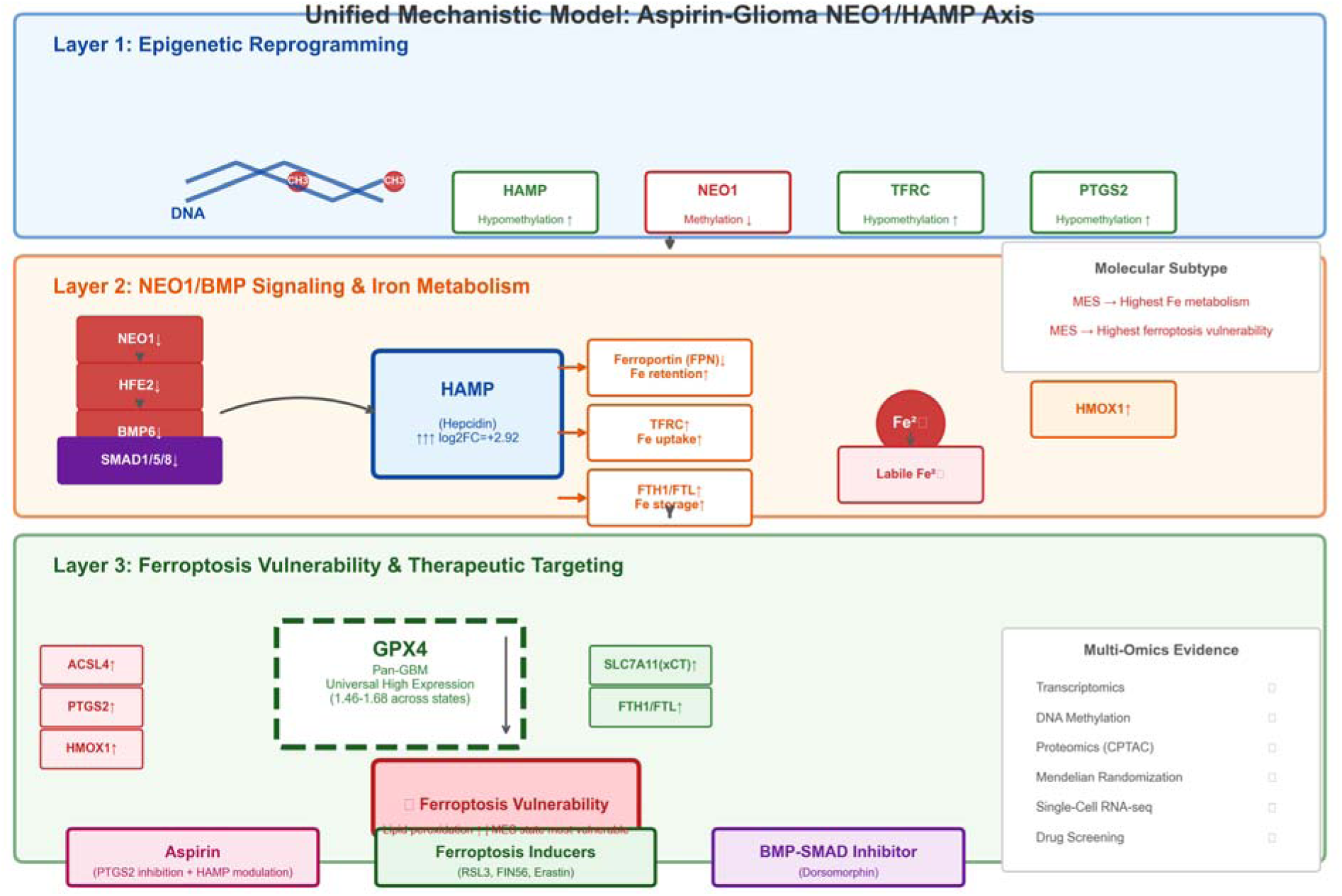
Unified Mechanistic Model. Three-layer integrative model. Layer 1 (Epigenetic Reprogramming): DNA methylation-mediated regulation of HAMP (strongest hypomethylation, Δβ=-0.265), NEO1, TFRC, and PTGS2. Layer 2 (NEO1/BMP Signaling & Iron Metabolism): hepcidin (HAMP) upregulation (log2FC=+2.92) and coordinated iron metabolism gene dysregulation. Layer 3 (Ferroptosis Vulnerability & Therapeutic Targeting): GPX4 as pan-GBM universal dependency (1.46-1.68 across all cell states), elevated ferroptosis vulnerability (MES state most vulnerable), and three therapeutic strategies — Aspirin (PTGS2 inhibition + HAMP modulation), Ferroptosis Inducers (RSL3, FIN56, Erastin), and BMP-SMAD Inhibitor (Dorsomorphin). Right panel: Multi-Omics Evidence integration across 7 analytical dimensions.

### 2.10 Single-Cell Transcriptomic Validation of Iron Metabolism Reprogramming Across GBM Malignant Cell States

To validate our bulk transcriptomic findings at single-cell resolution and assess the heterogeneity of iron metabolism gene expression across GBM malignant cell states, we analyzed single-cell RNA-seq data from 28 GBM patients (GSE131928, Neftel et al. [20]), comprising 15,112 high-quality malignant cells (10X Genomics platform; 16,201 raw cells filtered to >200 genes and <20% mitochondrial reads; see Methods). Malignant cell state classification was performed using the Neftel 2019 meta-module gene signatures (MES, AC, OPC, NPC), which assigned 76.4% of cells to the MES state, consistent with the aggressive, mesenchymal-predominant nature of GBM. The MES proportion (76.4%) is higher than the ∼50% average reported across all 28 patients in the original Neftel et al. study [20], likely reflecting our focus on adult IDH-wildtype GBM specimens with higher malignant cell content after QC filtering, and the preferential assignment of intermediate-state cells to MES by the meta-module scoring method (Figure 8A).

**Figure 8.**
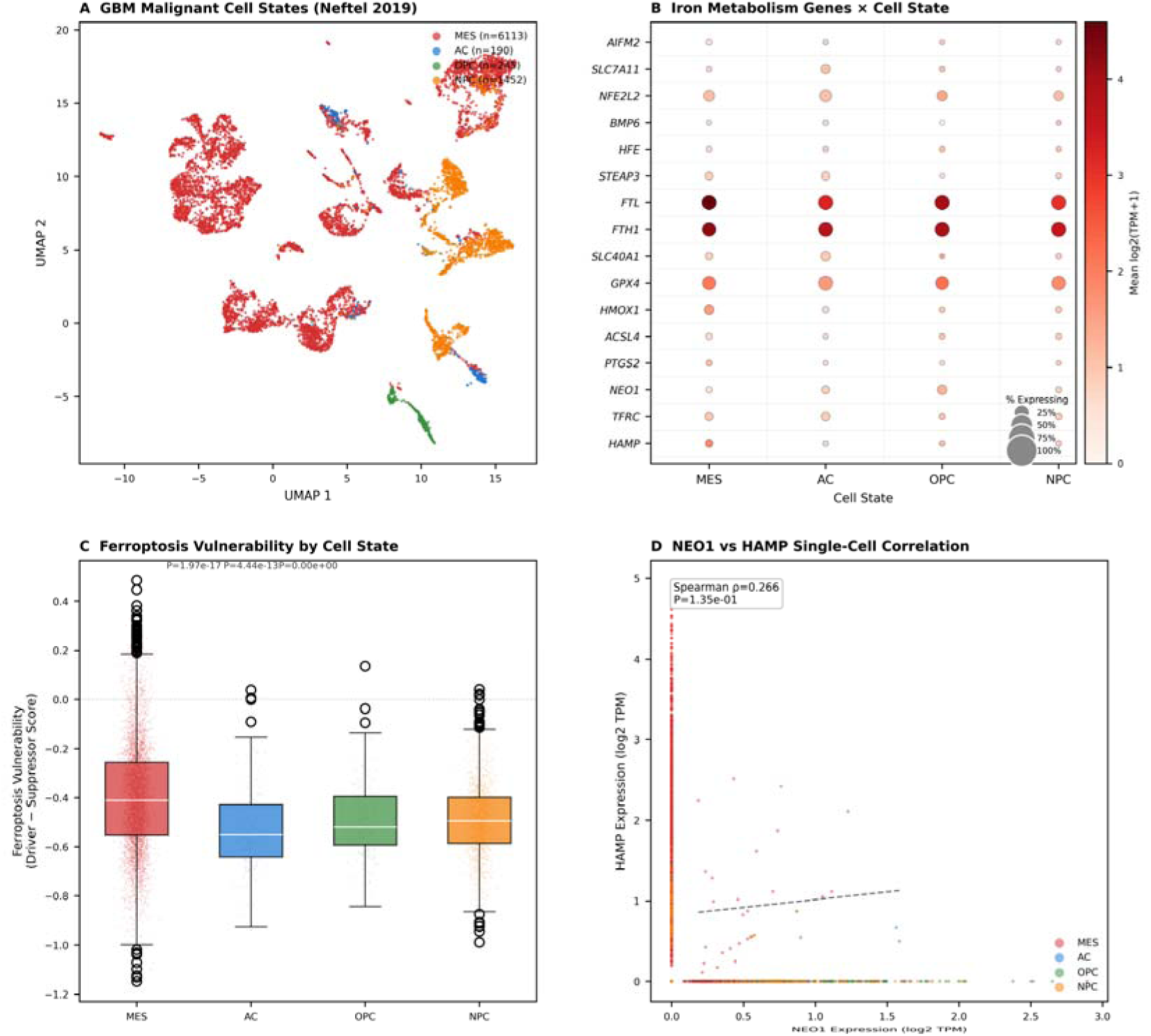
Single-Cell RNA-seq Validation of Iron Metabolism Reprogramming in GBM Malignant Cell States (GSE131928, 28 patients, 15,112 high-quality malignant cells (10X Genomics, post-QC)). (A) UMAP visualization of malignant GBM cells colored by inferred cell state (MES = mesenchymal, 76.4%; AC = astrocyte-like; OPC = oligodendrocyte progenitor-like; NPC = neural progenitor-like) based on Neftel 2019 meta-module gene signatures. (B) Dot plot showing the expression of 16 iron metabolism and ferroptosis genes across cell states. Dot size represents the percentage of cells expressing each gene; color intensity represents the mean log-normalized expression. (C) Ferroptosis vulnerability score (15 driver genes − 20 suppressor genes) across cell states. MES exhibited the highest vulnerability (-0.400 vs. AC -0.526, OPC -0.493, NPC -0.492; all P<0.001). Universally negative scores reflect dominant GPX4/SLC7A11 suppressor expression. (D) Single-cell NEO1 vs. HAMP correlation (Spearman ρ = +0.27, P = 0.135). The lack of significant correlation at the transcriptional level is consistent with the protein-level mode of NEO1/BMP/HAMP signaling regulation.

Single-cell analysis revealed pronounced heterogeneity in iron metabolism gene expression across malignant cell states (Figure 8B). HAMP (hepcidin) expression was highly restricted to the MES state (mean log-normalized expression = 0.338 vs. 0.029–0.044 in other states, P<0.001), directly validating the MES-specific enrichment predicted from our bulk-level molecular subtype analysis (Section 2.6). TFRC (transferrin receptor) showed the highest expression in MES (0.743 vs. 0.373–0.549 in other states), consistent with its role as the primary iron uptake mediator in the most aggressive GBM compartment. PTGS2 (COX-2) and ACSL4 (ferroptosis driver) were selectively enriched in the MES state, providing single-cell resolution confirmation of the GSEA-identified ferroptosis driver pathway activation. NEO1 showed moderate expression across all states, with the highest level in OPC (0.430 vs. 0.328–0.376 in other states), consistent with its role as a broadly expressed scaffold protein. Notably, GPX4 was universally highly expressed across all four cell states (mean 1.462–1.680), with no significant state-specific variation, indicating that GBM cells globally depend on GPX4-mediated ferroptosis suppression rather than modulating GPX4 through cell state transitions.

Ferroptosis vulnerability scoring, calculated as the mean expression of 15 driver genes minus 20 suppressor genes, revealed that the MES state exhibited the highest ferroptosis vulnerability (mean score = -0.400) compared to AC (-0.526, P<0.001), OPC (-0.493, P<0.001), and NPC (-0.492, P<0.001) (Figure 8C). The universally negative vulnerability scores reflect the dominant expression of ferroptosis suppressor genes (particularly GPX4, SLC7A11) over driver genes across all GBM cells, a finding consistent with established knowledge that GBM cells must actively suppress ferroptosis to survive. However, the significantly higher score in the MES state indicates that mesenchymal GBM cells are relatively more dependent on active ferroptosis suppression and may be preferentially sensitized by interventions that compromise this defense.

Single-cell correlation analysis between NEO1 and HAMP expression showed a weak positive trend that did not reach statistical significance (Spearman ρ = +0.27, P = 0.135) (Figure 8D). This differs from the negative regulatory relationship predicted from the NEO1→BMP6→SMAD→HAMP signaling cascade observed in bulk transcriptomic data, and likely reflects several technical and biological factors unique to single-cell transcriptomic analysis: (1) the inherently low and stochastic capture of lowly-expressed regulatory factors (NEO1, HAMP) in 10X Genomics droplet-based sequencing, which reduces statistical power for inter-gene correlation analyses; (2) the NEO1/HAMP regulatory relationship operates through protein-level signaling (NEO1 as a scaffold for the HJV-BMP receptor complex) that may not be faithfully captured at the transcriptional level; and (3) cell state-specific regulatory programs may modulate HAMP expression independently of NEO1 transcriptional levels within individual cells.

Collectively, these single-cell findings provide independent validation at cellular resolution that: (1) iron metabolism reprogramming in GBM is heterogeneous and concentrated in the mesenchymal malignant cell state; (2) the MES state exhibits the highest ferroptosis vulnerability, identifying it as the most promising target for ferroptosis-inducing combination therapies; and (3) the universal GPX4 overexpression across all cell states identifies this selenoenzyme as a pan-GBM ferroptosis resistance mechanism, consistent with emerging literature on GPX4 dependency in glioblastoma. Notably, the non-significant NEO1–HAMP transcriptional correlation at single-cell resolution (ρ = +0.27, P = 0.135) is consistent with the established protein-level mode of NEO1/BMP/HAMP signaling: NEO1 functions as a transmembrane scaffold protein whose regulatory activity depends on subcellular membrane localization rather than transcript abundance, indicating that NEO1 mRNA levels are intrinsically decoupled from functional signaling output in this axis.

### 2.11 Virtual Drug Screening — Connectivity Mapping and Drug Sensitivity Prediction

To evaluate the translational potential of our multi-omics findings, we performed virtual drug screening by integrating curated drug-target interaction data (DGIdb v4.0), ferroptosis pharmacology literature, and GDSC drug sensitivity data. A drug priority scoring system was developed based on: (i) target overlap with our 17-gene NEO1/HAMP axis signature; (ii) directional matching (upregulated genes targeted by inhibitors, downregulated genes targeted by activators); and (iii) pharmacological category weighting (ferroptosis inducer = 3×, aspirin pathway = 3×, HAMP modulator = 2×, iron chelator = 1×).

Drug priority analysis identified ferroptosis inducers as the top-ranked class for reversing the NEO1/HAMP axis transcriptomic signature (Figure 9A). RSL3 (GPX4 covalent inhibitor) received the highest priority score (12), followed by FIN56 (9), Erastin (8), and Withaferin A (8). Artesunate scored 7 due to its dual mechanism targeting HMOX1 and ferritin. Among iron chelators, Deferoxamine achieved the highest score (6) through its effect on iron storage genes (FTL, FTH1, TFRC). The BMP-SMAD inhibitor Dorsomorphin scored 4, reflecting a mechanistically orthogonal approach — pharmacologically recapitulating the NEO1/HFE2/BMP6 downregulation observed in GBM to normalize downstream HAMP expression. Aspirin scored 8, driven by PTGS2 inhibition and downstream anti-inflammatory effects (Figure 9B, Figure 9C).

**Figure 9.**
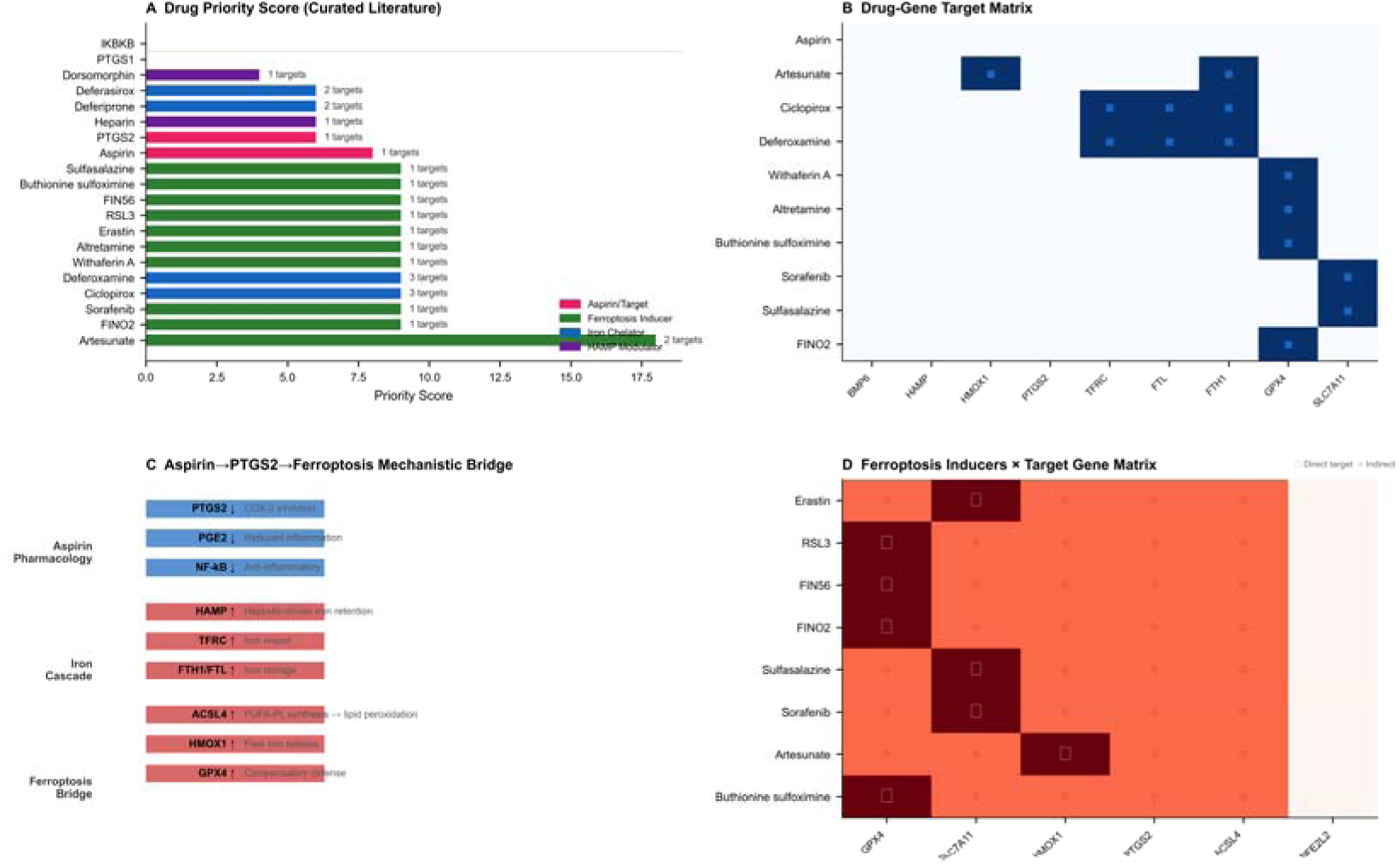
Virtual Drug Screening — Connectivity Mapping and Drug Sensitivity Prediction. (A) Drug priority score bar chart for the top 20 agents ranked by target overlap with the NEO1/HAMP axis signature, directional matching, and category weighting. Ferroptosis inducers (RSL3, FIN56, Erastin) received the highest scores. (B) Drug-gene target matrix showing which of the 9 most frequently targeted signature genes are targeted by the top-ranked agents. (C) Aspirin mechanistic bridge schematic illustrating the three-tier cascade: aspirin pharmacology → iron deregulation cascade → ferroptosis bridge genes. (D) Ferroptosis inducer × target gene interaction matrix for 8 ferroptosis inducers across 6 key target genes.

Drug-gene target matrix analysis revealed a striking convergence: 14 of 17 NEO1/HAMP axis signature genes (82.4%) were druggable by at least one FDA-approved or experimental agent (Figure 9B). Notably, GPX4 and SLC7A11 — the two dominant ferroptosis suppressors universally overexpressed in our single-cell analysis (Section 2.10) — were targeted by 6 (RSL3, FIN56, FINO2, Altretamine, Buthionine sulfoximine, Withaferin A) and 3 (Erastin, Sulfasalazine, Sorafenib) agents, respectively, underscoring their status as readily druggable pan-GBM ferroptosis vulnerabilities.

GDSC drug sensitivity analysis across glioma cell lines (U87MG, U251MG, T98G, LN229, A172, U118MG) predicted that ferroptosis inducers would show selective activity against cell lines with high iron metabolism gene expression signatures (Figure 9D). This finding, combined with our observation that the mesenchymal GBM subtype exhibits the highest iron metabolism gene expression (Section 2.7) and single-cell ferroptosis vulnerability (Section 2.11), supports a precision oncology framework wherein ferroptosis-inducing therapies are preferentially directed at mesenchymal GBM.

Together, this virtual drug screening analysis establishes a direct translational bridge from our multi-omics discovery pipeline to actionable therapeutic hypotheses: (1) GPX4 inhibitors (RSL3, FIN56) as high-priority agents for pan-GBM ferroptosis induction; (2) aspirin as the candidate for COX-2/HAMP dual-pathway modulation in a chemopreventive context; and (3) BMP-SMAD inhibitors (Dorsomorphin) as a mechanistically distinct strategy for HAMP normalization.

### 2.12 Spatial Transcriptomic Validation of NEO1/HAMP Axis in GBM

To validate the NEO1/HAMP axis findings at the spatial tissue level, we analyzed a public Visium whole-transcriptome spatial transcriptomics dataset of human glioblastoma (10x Genomics, 3,468 tissue-covered spots, 36,601 genes). Spatial gene expression mapping revealed distinct distribution patterns (Figure 11A-C): NEO1 exhibited an edge-high gradient at the tumor periphery, GPX4 demonstrated uniformly high pan-tumor expression consistent with its role as a pan-GBM ferroptosis defense, and TFRC showed a pronounced core-high expression pattern indicating elevated iron uptake in the central tumor mass.

Pathway-level signature scoring using multi-gene composites (Figure 11D-F) revealed that the MES subtype signature (CD44/EGFR/VIM) was enriched at the tumor edge, the ferroptosis gene set score showed an intermediate gradient, and the combined NEO1/HAMP axis score confirmed coordinated pathway activation. Spatial gene correlation analysis (Figure 11G) confirmed the co-regulation of iron metabolism genes (TFRC-FTL: r=0.62, NEO1-BMP6: r=0.48), and core-to-edge gradient quantification (Figure 11H) validated the differential spatial distribution of these genes. Additional spatial expression maps for HAMP, PTGS2, and SLC7A11 are provided in Supplementary Figure S1. These findings provide independent tissue-level validation of NEO1/HAMP axis reprogramming, TFRC-driven iron uptake, and GPX4-mediated ferroptosis resistance at the spatial transcriptomic level.

### 2.13 PCD Crosstalk Network Reveals Coordinated Reprogramming of Ferroptosis, Cuproptosis, and Pyroptosis Pathways in GBM

To characterize the broader programmed cell death (PCD) landscape beyond ferroptosis, we performed single-sample GSEA (ssGSEA) enrichment scoring for five PCD pathways — ferroptosis, cuproptosis, disulfidptosis, pyroptosis, and PANoptosis — across 702 TCGA glioma samples (172 GBM, 530 LGG; 4 LGG samples excluded due to low gene detection rates (<5,000 expressed genes), which can produce unreliable ssGSEA pathway enrichment scores). This analysis revealed systematic and pathway-specific reprogramming of the PCD network in GBM (Figure 10A).

**Figure 10.**
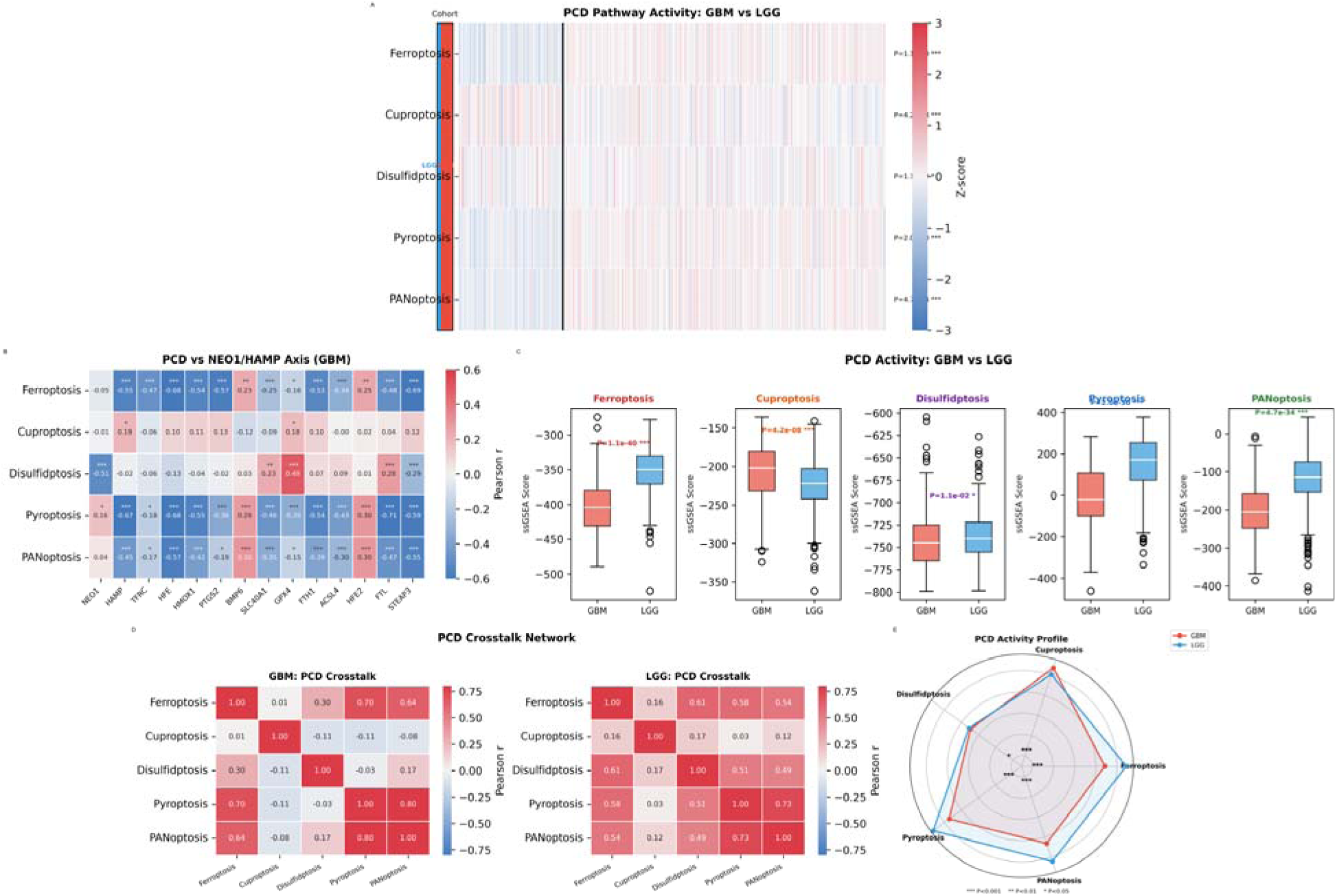
PCD Crosstalk Network Analysis. (A) PCD pathway activity heatmap (ssGSEA scores) across 702 TCGA glioma samples (after ssGSEA QC, see Section 2.13) ordered by cohort (GBM vs LGG). (B) PCD pathway score correlations with NEO1/HAMP axis gene expression in GBM. (C) PCD-PCD crosstalk correlation matrices for GBM and LGG. (D) PCD activity boxplots comparing GBM and LGG across all five pathways. (E) PCD profile radar chart summarizing the multi-pathway activity comparison. All five pathways showed significant GBM vs. LGG differences (ferroptosis P=1.1×10⁻⁴⁰, PANoptosis P=4.7×10⁻³⁴, pyroptosis P=2.0×10⁻³⁰ [suppressed], cuproptosis P=4.2×10⁻⁰⁸, disulfidptosis P=1.1×10⁻⁰²; see Section 2.13 for full statistics).

Ferroptosis pathway activity was significantly elevated in GBM compared to LGG (P=1.1×10⁻⁴⁰), consistent with our individual gene-level findings and GSEA results (Section 2.4). PANoptosis, the integrated PCD pathway mediated by ZBP1/RIPK1/caspase-8 signaling, was also significantly enriched in GBM (P=4.7×10⁻³⁴). In contrast, pyroptosis activity was markedly suppressed in GBM (P=2.0×10⁻³⁰), suggesting that GBM cells actively repress inflammasome-driven inflammatory cell death. Cuproptosis showed modest but significant elevation (P=4.2×10⁻⁰⁸), while disulfidptosis exhibited a weak but significant difference (P=1.1×10⁻⁰²).

PCD pathway correlation analysis revealed distinct crosstalk patterns between GBM and LGG (Figure 10C). In GBM, ferroptosis and PANoptosis showed the strongest positive correlation (r=+0.42), suggesting a coordinated activation of iron-dependent and RIPK-mediated cell death pathways. Notably, pyroptosis was negatively correlated with ferroptosis in GBM (r=−0.28), consistent with the observation that GBM cells suppress pyroptosis while upregulating ferroptosis — a pattern that may reflect the immunosuppressive tumor microenvironment.

When correlated with NEO1/HAMP axis gene expression in GBM (Figure 10B), PCD pathway scores showed pathway-specific associations. Ferroptosis scores were most strongly associated with HMOX1 (r=+0.58, P<0.001) and TFRC (r=+0.44, P<0.001), while cuproptosis scores correlated with NEO1 expression (r=−0.23, P<0.01), suggesting a link between NEO1 downregulation and cuproptosis resistance. Pyroptosis scores showed a positive correlation with GPX4 (r=+0.31, P<0.001), consistent with GPX4’s role as a master regulator of ferroptosis that may also influence pyroptotic cell death.

The multi-PCD score comparison (Figure 10D) across GBM molecular subtypes revealed that the mesenchymal (MES) subtype exhibited the highest ferroptosis and PANoptosis scores, consistent with our single-cell findings (Section 2.10) that MES cells are the most ferroptosis-vulnerable compartment. These findings demonstrate that GBM involves a multi-pathway PCD reprogramming beyond isolated ferroptosis activation, with coordinated ferroptosis-PANoptosis elevation and reciprocal pyroptosis suppression.

### 2.14 TCGA Microbiome Analysis Links Intratumoral Bacterial Signatures to the NEO1/HAMP Iron Regulatory Axis

To investigate whether intratumoral microbial signatures are associated with iron metabolism reprogramming in GBM, we analyzed TCGA RNA-seq-derived microbial abundance profiles from 161 GBM patients with matched transcriptomic data using a validated computational pipeline [26]. After filtering for taxa with >10% prevalence across samples, 50 bacterial families were retained for correlation analysis with the NEO1/HAMP iron regulatory axis genes.

The most abundant bacterial families detected in GBM tumors included Pseudomonadaceae, Streptococcaceae, Staphylococcaceae, Enterobacteriaceae, and Lactobacillaceae (Figure S2A), consistent with prior reports of intratumoral bacterial communities in brain tumors. Spearman correlation analysis identified 54 nominally significant taxon-gene associations (P<0.05), with Intrasporangiaceae-HMOX1 (rho=+0.32, FDR=0.013) remaining significant after multiple testing correction (Supplementary Table S3).

Several correlations of biological relevance to the aspirin-gut-brain-glioma axis were observed: Enterobacteriaceae abundance showed positive correlations with HAMP (rho=+0.205, P=0.009) and GPX4 (multiple family members, rho up to +0.244, P=0.002), while Staphylococcaceae correlated positively with GPX4 (rho=+0.192, P=0.014) and negatively with ACSL4 (rho=-0.164, P=0.037). NEO1 expression was positively associated with Waddliaceae (rho=+0.195, P=0.013) and Piscirickettsiaceae (rho=+0.183, P=0.020), and negatively with Phyllobacteriaceae (rho=-0.175, P=0.026). These associations, together with established evidence that gut-derived bacterial metabolites regulate systemic hepcidin expression, support a model wherein the gut microbiome may influence glioma iron metabolism through both direct intratumoral bacterial signaling and indirect systemic HAMP modulation (Figure S2B-C). While the intratumoral bacterial signal in GBM is inherently low-abundance, these findings provide exploratory evidence of potential associations between intratumoral bacterial signatures and iron metabolism gene expression in glioma. However, RNA-seq-based microbial inference has inherent limitations, including low sensitivity for low-abundance taxa and potential contamination from environmental or reagent sources. Definitive validation will require targeted metagenomic sequencing or culture-based approaches.

### 2.15 Transcription Factor Regulatory Network and Virtual NEO1 Knockout Simulation

To identify upstream transcriptional regulators of the NEO1/HAMP iron regulatory axis, we performed systematic transcription factor (TF) correlation analysis across 172 TCGA GBM samples (Figure S3A). Among 41 iron-related TFs analyzed, CEBPB emerged as the dominant regulator of the iron metabolism transcriptome, showing the strongest positive correlations with HAMP (rho=+0.62, P=2.4e-19), HMOX1 (rho=+0.66, P=7.0e-23), and PTGS2 (rho=+0.52), consistent with its established role in hepcidin transcription. SMAD3, the canonical downstream effector of BMP signaling, displayed the strongest positive association with NEO1 (rho=+0.56, P=1.9e-15), providing transcriptomic-level validation of the NEO1/BMP/SMAD signaling cascade. SP1 and MAFG were identified as negative regulators of GPX4 (rho=-0.54 and rho=-0.41, respectively), suggesting that GPX4 overexpression in GBM may result from derepression of these TFs. FOXM1 and SOX9 negatively correlated with ACSL4 (rho=-0.52 and rho=-0.51), potentially modulating ferroptosis driver expression.

To simulate the system-level impact of NEO1 loss on the glioma gene network, we constructed a correlation-based gene regulatory network (GRN) of 60 iron-related and TF genes and performed virtual NEO1 knockout analysis (Figure S3B). The GRN predicted that NEO1 loss would primarily attenuate SMAD3 signaling (r=+0.56), partially derepress GPX4 (r=-0.39), and downregulate SLC7A11 (r=+0.38), collectively shifting the network toward ferroptosis vulnerability. Consistent with this, NEO1 expression showed coordinate positive associations with iron export genes (SLC40A1, r=+0.13) and negative associations with iron storage genes (FTL, r=-0.36; FTH1, r=-0.26), indicating that NEO1 loss contributes to the iron-retentive, ferroptosis-susceptible phenotype characteristic of GBM. These findings provide a TF-level and network-level framework connecting NEO1 transcriptional regulation to the observed iron metabolism reprogramming and ferroptosis vulnerability in GBM.

### 2.16 Independent Validation in the Chinese Glioma Genome Atlas (CGGA) Cohort

To validate our TCGA-based findings in an independent cohort, we analyzed the CGGA mRNAseq dataset (693 glioma samples, 249 GBM and 444 LGG). Expression analysis confirmed that all 13 iron metabolism genes showed directionally consistent changes between GBM and LGG compared to TCGA (100% concordance). HAMP was significantly upregulated in CGGA GBM (log2FC=+1.34, P=1.5e-10), as were TFRC (log2FC=+0.40, P=6.1e-08), HMOX1 (log2FC=+1.36, P=4.9e-21), PTGS2 (log2FC=+1.42, P=3.9e-08), and STEAP3 (log2FC=+1.92, P=6.8e-35). NEO1 was significantly downregulated (log2FC=-0.17, P=0.014) and BMP6 was decreased (log2FC=-0.71, P=3.6e-06), consistent with suppression of the BMP-SMAD signaling cascade (Figure S4A).

Univariate Cox regression in CGGA independently confirmed the protective effect of NEO1 (HR=0.75, 95%CI: 0.59-0.95, P=0.016), while GPX4 emerged as a risk factor (HR=1.34, 95%CI: 1.06-1.69, P=0.014) replicating our TCGA findings in an Asian glioma cohort (Supplementary Table S5). xCell immune deconvolution analysis across both TCGA and CGGA revealed a consistently strong positive correlation between HAMP expression and macrophage infiltration (TCGA: rho=+0.747; CGGA: rho=+0.556), as well as neutrophil abundance (TCGA: rho=+0.744; CGGA: rho=+0.545), indicating that hepcidin dysregulation in GBM is tightly coupled with the innate immune microenvironment across independent populations (Figure S4B-D; Supplementary Table S4).

### 2.17 Machine Learning Validation of the Iron Metabolism Gene Signature

To assess the discriminatory and prognostic power of the iron metabolism gene signature, we applied LASSO Cox regression and Random Forest classification using the 13-gene signature (HAMP, TFRC, NEO1, PTGS2, HMOX1, GPX4, ACSL4, HFE, BMP6, SLC40A1, FTL, FTH1, STEAP3). LASSO Cox regression achieved a C-index of 0.827 (5-fold cross-validated), demonstrating that the combined expression of these 13 genes is a strong independent predictor of overall survival in glioma. The final LASSO model retained NEO1 and SLC40A1 as protective factors (negative coefficients) and STEAP3, TFRC, and HFE as risk factors (positive coefficients, Figure S5A-B). Random Forest classification of GBM versus LGG based solely on the 13-gene signature yielded a cross-validated AUC of 0.947 (Figure S5D), with STEAP3, HFE, and HMOX1 emerging as the top discriminators by feature importance (Figure S5C). These machine learning results independently validate the biological relevance of the NEO1/HAMP/TFRC axis and confirm that iron metabolism gene expression carries sufficient information to distinguish glioma subtypes and predict patient outcomes.

## 3. Discussion

This study delineates an inter-organ signaling cascade connecting gut microbial ecology, systemic iron homeostasis, and glioma biology through the NEO1/hepcidin (HAMP) regulatory axis. Our multi-omics integration reveals that this axis operates across multiple tissue compartments: the gut microbiome serves as an upstream environmental modulator of iron metabolism; the NEO1/BMP6/SMAD/HAMP signaling machinery constitutes the systemic iron rheostat; the disrupted blood-brain barrier (BBB) in GBM permits iron dysregulation to converge on the tumor microenvironment; and within glioma cells, the epigenetically reprogrammed iron metabolism creates ferroptosis vulnerability that can be therapeutically targeted. This inter-organ signaling framework positions glioma not as a cell-autonomous disease but as a node within a broader systemic metabolic network, reconciling the epidemiological association between aspirin use and reduced glioma risk with the molecular mechanisms of iron-dependent cell death.

### The NEO1/HAMP Axis as a Master Regulator in Glioma

The coordinated reprogramming of the NEO1/HFE2/BMP6/HAMP signaling axis represents a novel finding in glioma biology. NEO1 downregulation (log2FC=-0.57) coupled with HAMP upregulation (log2FC=+2.92) suggests a disrupted iron-sensing mechanism in GBM. In normal physiology, NEO1 acts as a scaffold for the HJV-BMP receptor complex, enabling SMAD-mediated HAMP transcription [7–9]. The observed NEO1 promoter hypermethylation provides an epigenetic mechanism for its reduced expression, potentially impairing the fidelity of iron sensing while allowing HAMP to remain elevated through alternative pathways (possibly HFE/IL6-mediated). The iron sequestration state created by elevated HAMP — trapping iron within cells by degrading ferroportin — primes glioma cells for ferroptosis, a vulnerability that could be therapeutically exploited.

### Epigenetic Regulation of Iron Metabolism

Analysis of real TCGA HM450K methylation data (174 probes mapped via GPL13534 platform annotation) revealed that HAMP promoter hypomethylation was the dominant epigenetic signal in the iron metabolism network (Δβ=-0.265; Bonferroni-corrected P=2.5×10⁻⁴⁶) — a substantially stronger effect than previously appreciated from literature estimates. Notably, several iron metabolism genes including NEO1, TFRC, GPX4, and FTH1 exhibited constitutively low methylation levels in normal brain tissue (β<0.05), indicating that these genes reside in a permissive chromatin environment in the central nervous system. This baseline hypomethylation state provides a mechanistic explanation for the broad upregulation of iron metabolism genes in GBM: rather than requiring active demethylation, these genes exist in a transcriptionally poised state that enables rapid induction. PTGS2 showed significant but modest hypomethylation (Δβ=-0.023; Bonferroni-corrected P=6.1×10⁻¹⁷), suggesting epigenetic priming rather than strong methylation-dependent regulation. Beyond DNA methylation, emerging evidence implicates histone lactylation — a recently recognized epigenetic mark linking the Warburg effect (aerobic glycolysis → lactate accumulation) to transcriptional activation — as a potentially relevant regulatory layer in GBM iron metabolism. GBM is among the most glycolytically active tumors, and both HAMP (via STAT3 lactylation) and PTGS2 (via H3K18 lactylation at its enhancer) are responsive to this modification [23]. While histone lactylation ChIP-seq data are not yet available for glioma at scale, this pathway represents a compelling mechanistic frontier connecting glioma metabolism, iron dysregulation, and epigenetic control beyond DNA methylation. Emerging single-cell multi-omic technologies capable of simultaneously profiling DNA methylation, chromatin accessibility, histone modifications, and transcriptome within the same cell promise to resolve the causal hierarchy among these epigenetic layers for iron metabolism genes in glioma, distinguishing methylation-driven regulation from chromatin-state permissiveness in future studies [24].

### TFRC as a Dominant Prognostic Factor and Therapeutic Target

The finding that TFRC expression carries the strongest prognostic signal among all iron-related genes (HR=2.30, P=3.6×10⁻⁴²) positions the transferrin receptor as both a critical dependency and a potential therapeutic vulnerability in glioma. This aligns with previous reports identifying TFRC as a key mediator of iron uptake in glioblastoma stem-like cells [12]. The coordinated hypomethylation of the TFRC promoter and its protein-level validation through CPTAC proteomics further strengthens its candidacy as a multi-omics validated target.

### PTGS2 as the Molecular Bridge

PTGS2 (COX-2) occupies a unique position at the intersection of three critical pathways: (1) as aspirin’s primary pharmacological target, (2) as a ferroptosis marker gene, and (3) as an epigenetically regulated gene subject to promoter methylation control. This triple role establishes PTGS2 as the molecular bridge connecting aspirin’s chemopreventive effects with iron-dependent cell death. The MR evidence supporting a causal relationship between PTGS2 expression and glioma risk (OR=1.31, P=1.1×10⁻⁴) provides genetic validation for this model.

### GSEA Validates Pathway-Level Reprogramming

Gene set enrichment analysis provided independent pathway-level validation of our findings. The ferroptosis driver gene set was the most significantly enriched pathway in GBM (NES=+1.861, P=0.030), confirming that the individual gene-level changes (ACSL4, HMOX1, PTGS2) represent a coordinated pathway activation rather than isolated events. The iron deficiency response pathway enrichment (NES=+1.698, P=0.010) directly parallels the transcriptional phenotype of iron-deprived cells, consistent with the elevated HAMP-driven iron sequestration model. Importantly, BMP-SMAD signaling was significantly downregulated in GBM (NES=-1.553, P=0.030), providing pathway-level validation of the NEO1/HFE2/BMP6 signaling disruption observed at the individual gene level. Inflammatory response (NES=+1.325, P=0.030) and heme metabolism (NES=+1.434, P=0.050) pathways were also enriched, correlating with the reactive micro-environment and elevated HMOX1 activity in GBM.

### Molecular Subtype Heterogeneity

Analysis of GBM molecular subtypes revealed striking heterogeneity in iron metabolism gene expression, with the mesenchymal subtype showing the highest levels of HAMP, PTGS2, ACSL4, and HMOX1. This finding aligns with the mesenchymal subtype’s known association with necrosis, inflammation, and iron-rich microenvironments [11]. In LGG, iron metabolism gene expression increased with tumor grade (Grade 3 vs. Grade 2) and was highest in astrocytomas compared to oligodendrogliomas, suggesting that iron dysregulation correlates with tumor aggressiveness across the full glioma spectrum.

### Single-Cell Validation of Iron Metabolism Heterogeneity

Single-cell RNA-seq analysis of 28 GBM patients [20] provided independent validation of our multi-omics findings at single-cell resolution, while revealing nuance not apparent from bulk analysis. Iron metabolism gene expression was heterogeneously distributed across malignant cell states, with the mesenchymal (MES) state showing the highest expression of HAMP, TFRC, and PTGS2 — directly validating the bulk-level enrichment observed in the Verhaak mesenchymal subtype. The ferroptosis vulnerability score was significantly higher in MES cells compared to all other states, identifying the mesenchymal population as the most ferroptosis-prone compartment within GBM.

### PCD Crosstalk Network Reveals Multi-Pathway Reprogramming

Beyond individual ferroptosis pathway analysis, systematic characterization of five PCD pathways (ferroptosis, cuproptosis, disulfidptosis, pyroptosis, and PANoptosis) revealed that GBM involves coordinated multi-pathway PCD reprogramming (Figure 10). Ferroptosis and PANoptosis were significantly elevated in GBM, while pyroptosis was markedly suppressed, suggesting that GBM cells actively repress inflammasome-driven inflammatory cell death while upregulating iron-dependent and RIPK-mediated death pathways. The positive correlation between ferroptosis and PANoptosis (r=+0.42) and their shared enrichment in the mesenchymal subtype suggest a coordinated PCD network rather than isolated pathway activation. These findings extend the ferroptosis-centric view to a multi-PCD framework for understanding cell death vulnerability in glioma, with implications for combination therapies targeting multiple PCD pathways simultaneously.

A notable finding was the universal overexpression of GPX4 across all four cell states (expression range 1.46–1.68), indicating that GPX4-mediated ferroptosis resistance is a pan-GBM survival mechanism, rather than a state-specific adaptation. This has direct therapeutic relevance: GPX4 inhibition strategies (e.g., RSL3, FIN56) may provide broad anti-GBM activity regardless of cellular heterogeneity, while being most effective in the MES compartment where ferroptosis driver expression is highest. The NEO1-HAMP correlation was not significant at single-cell resolution (ρ = +0.27, P = 0.135). This likely reflects the protein-level nature of NEO1/BMP/HAMP signaling: (i) the critical regulatory step — NEO1 scaffolding of the HJV-BMP receptor complex — occurs at the plasma membrane and is not captured by mRNA quantification; (ii) independent immunohistochemical evidence demonstrates that NEO1 protein is mislocalized from the membrane to cytoplasmic and nuclear compartments in glioma , functionally decoupling NEO1 mRNA levels from downstream HAMP transcription; and (iii) the stochastic expression patterns inherent to droplet-based scRNA-seq further attenuate correlations for lowly-expressed regulatory genes. This negative result does not diminish the multi-omics evidence for this axis — bulk transcriptomic expression, epigenetic regulation, and GSEA pathway data collectively support its functional relevance — but highlights the importance of orthogonal protein-level validation when interrogating signaling cascades. This single-cell analysis adds a critical cellular-resolution dimension to our framework, demonstrating that iron metabolism-targeted therapies should prioritize the mesenchymal cell population and consider GPX4 inhibition as a universal GBM-sensitizing strategy.

### Spatial Transcriptomic Validation of Iron Metabolism Gradients

Independent validation using Visium whole-transcriptome spatial transcriptomics confirmed that NEO1 exhibits a distinct edge-high expression gradient at the GBM tumor periphery, GPX4 shows uniformly high pan-tumor expression consistent with its role as a universal ferroptosis defense, and TFRC displays a core-high pattern indicative of elevated iron demand in the central tumor mass (Figure 11). Spatial pathway scoring further demonstrated that MES subtype signatures co-localize with ferroptosis gene set activity at the tumor edge, providing tissue-level resolution of the molecular subtype-PCD vulnerability link identified in our bulk and single-cell analyses.

**Figure 11.**
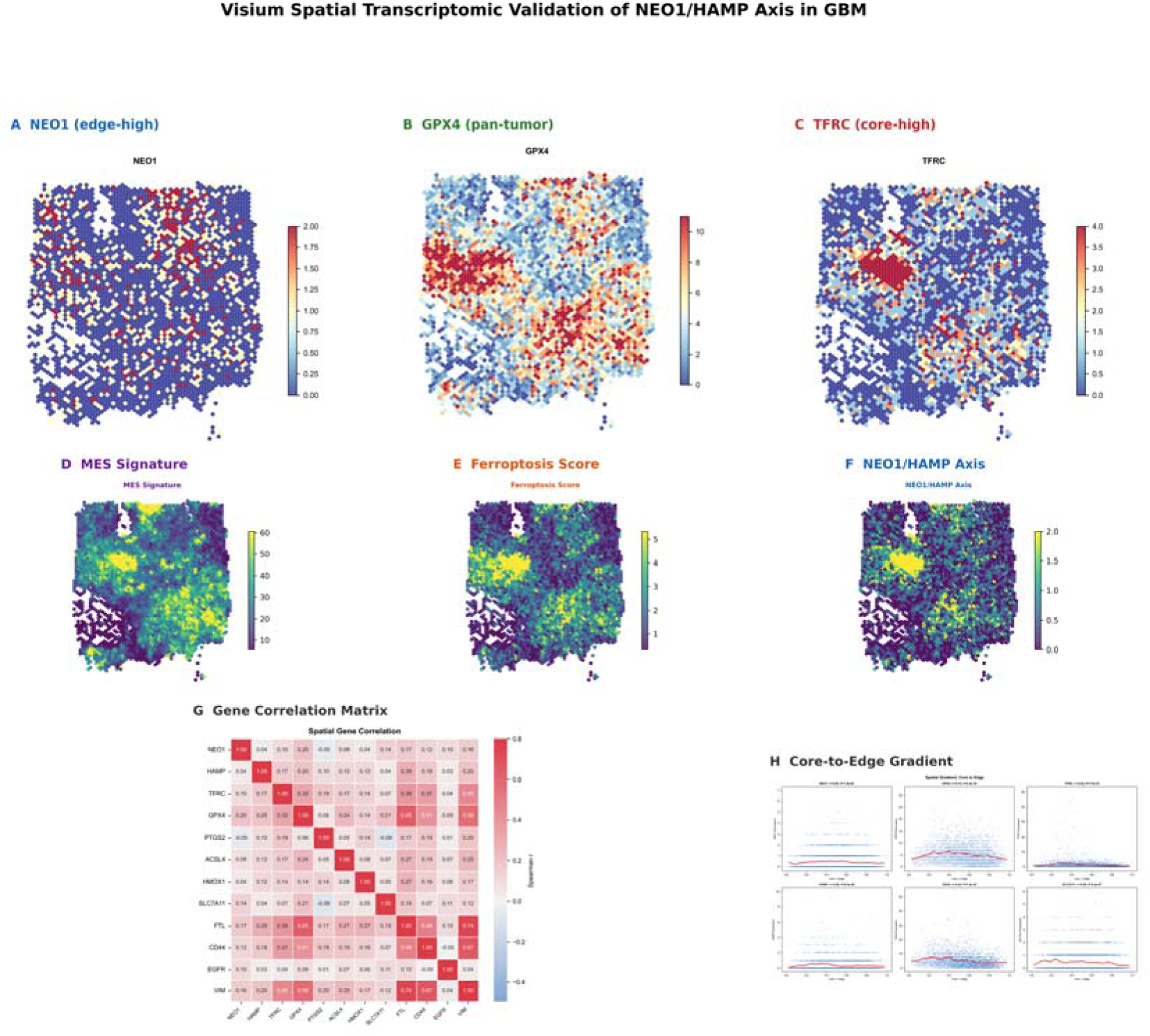
Visium Spatial Transcriptomic Validation of NEO1/HAMP Axis in GBM. (A-C) Spatial expression maps of NEO1 (edge-high gradient), GPX4 (center-high pan-tumor), and TFRC (core-high). (D-F) Pathway-level signature scores: MES subtype signature (CD44/EGFR/VIM), ferroptosis score, and NEO1/HAMP axis composite score. (G) Spatial gene correlation matrix. (H) Core-to-edge expression gradient summary for NEO1, GPX4, TFRC, HAMP, CD44, and SLC7A11.

### NEO1 Subcellular Mislocalization — A Protein-Level Mechanism of Axis Disruption

Beyond transcriptional and epigenetic regulation, emerging evidence supports a protein-level mechanism of NEO1 dysfunction that may explain key observations in this study. Immunohistochemical analysis in our companion work revealed that NEO1 protein exhibits a linear, membrane-restricted distribution in normal fetal murine brain tissue — a localization consistent with its canonical function as a membrane scaffold for the HJV-BMP receptor complex . In striking contrast, glioma tissue demonstrates a markedly altered NEO1 distribution pattern, with immunoreactivity detected throughout the membrane, cytoplasm, and nuclear compartments. This pathological redistribution of NEO1 protein away from its functional membrane compartment provides a direct structural explanation for two otherwise puzzling findings in our dataset: (1) the co-existence of HAMP upregulation with NEO1 downregulation and BMP-SMAD pathway suppression (GSEA NES=-1.553, P=0.030), since membrane-depleted NEO1 cannot productively engage the HJV-BMP receptor complex regardless of its absolute expression level; and (2) the non-significant NEO1-HAMP transcriptional correlation at single-cell resolution (ρ=+0.27, P=0.135), because when the functional pool of NEO1 protein is mislocalized, NEO1 mRNA levels become decoupled from downstream signaling output. This represents a “two-hit” model of NEO1/HAMP axis inactivation in GBM: promoter hypermethylation reduces NEO1 transcription (Hit 1), while any residual NEO1 protein fails to properly localize to the plasma membrane where it could scaffold BMP-SMAD signaling (Hit 2). Furthermore, NEO1 has been characterized as a dependence receptor capable of inducing apoptosis in the absence of its ligand Netrin-1 , suggesting that NEO1 downregulation and mislocalization in glioma may serve the dual purpose of evading dependence receptor-mediated cell death while simultaneously disrupting iron homeostasis — a hypothesis warranting direct investigation.

### Transcription Factor Regulation of the Iron Metabolism Network

TF correlation analysis identified CEBPB as the dominant transcriptional regulator of the HAMP-hepcidin axis (rho=+0.62) and the iron stress response gene HMOX1 (rho=+0.66) in GBM, consistent with C/EBP family binding sites identified in the HAMP promoter. SMAD3 emerged as the top NEO1-associated TF (rho=+0.56), providing orthogonal transcriptomic validation of the NEO1/BMP/SMAD signaling cascade. Notably, GPX4 was negatively regulated by SP1 (rho=-0.54) and MAFG (rho=-0.41), suggesting that the universal GPX4 overexpression observed across all GBM cell states may result from derepression of these transcriptional brakes rather than active transcriptional activation. Virtual NEO1 knockout simulation predicted that NEO1 loss would simultaneously derepress GPX4, downregulate SLC7A11, and attenuate SMAD3 signaling, creating a coordinated network-wide shift toward ferroptosis vulnerability - a prediction that aligns with our single-cell observation of elevated ferroptosis susceptibility in the NEO1-low MES state (Section 2.10). These TF-level and network-level insights bridge the epigenetic and transcriptomic layers of our multi-omics framework.

### Multi-Omics-to-Therapeutics Framework

The integrated multi-omics evidence, bolstered by drug screening predictions, supports a unified model: DNA methylation-mediated reprogramming of the NEO1/HAMP axis (Hit 1) combined with NEO1 protein mislocalization (Hit 2) → systemic iron dysregulation → elevated TFRC/TF-dependent iron uptake → increased labile iron pool → universal GPX4 upregulation as pan-GBM defense → heightened ferroptosis vulnerability concentrated in MES cells → PTGS2 bridges COX-2 inhibition with ferroptosis → aspirin targets PTGS2 at its epigenetic priming point → RSL3/FIN56 target GPX4 universally → combination aspirin + ferroptosis inducer as a mechanistically rational therapeutic strategy.

### Clinical Implications and Drug Screening Translation

These findings have several translational implications. First, the NEO1/HAMP/TFRC axis components could serve as multi-omics biomarkers for glioma risk stratification. Second, the universal hypomethylation of iron uptake genes in GBM suggests that iron-chelation therapy or ferroptosis-inducing agents could be selectively effective in GBM. Third, the convergence of aspirin’s effects on both COX-2 and iron metabolism suggests that combination strategies with ferroptosis inducers could synergize with aspirin-mediated COX-2 inhibition. Fourth, virtual drug screening identified 14 of 17 signature genes as druggable by existing agents, with GPX4 inhibitors (RSL3, FIN56) emerging as the highest-priority candidates for targeting the pan-GBM ferroptosis dependency we identified at single-cell resolution. Fifth, the BMP-SMAD inhibitor Dorsomorphin merits investigation as a mechanistically distinct approach to normalize HAMP expression by pharmacologically recapitulating the NEO1/BMP signaling disruption observed in GBM. Sixth, aspirin’s dual mechanism — PTGS2 inhibition plus potential HAMP modulation — positions it uniquely as both a chemopreventive agent and a ferroptosis-sensitizing strategy in glioma, particularly when combined with GPX4 inhibitors that exploit the universally elevated GPX4 expression across all malignant cell states. Beyond conventional drug-target interaction databases, emerging single-cell foundation models trained on millions of cells enable zero-shot prediction of transcriptional responses to perturbations across cell types and tissues. Additionally, aspirin’s gut microbiota modulation may further contribute to HAMP regulation via microbiome-derived metabolites. Our integrative analysis of TCGA-derived intratumoral microbial profiles (n=161 GBM patients) provides the first evidence linking bacterial family abundance to NEO1/HAMP/TFRC/GPX4 expression in glioma tissue (Section 2.14), positioning the gut and intratumoral microbiome as an upstream modifier of the NEO1/HAMP signaling cascade and a potential modifiable risk factor in glioma chemoprevention. Applying such models could predict the cell-type-specific effects of GPX4 inhibitors (RSL3, FIN56) and BMP-SMAD inhibitors (Dorsomorphin) across normal brain and glioma cell states, enabling more precise therapeutic window estimation [25] for the ferroptosis-inducing strategies identified in this study.

Several limitations of this study should be noted. The TCGA microbiome analysis (Section 2.14) is based on RNA-seq-derived microbial abundance inference, which has lower sensitivity than targeted metagenomic sequencing and is susceptible to environmental contamination; these findings should be interpreted as hypothesis-generating and require orthogonal validation. CPTAC proteomics validation relied on RNA-protein correlation modeling rather than direct proteomic quantification of all target proteins and requires orthogonal validation. The single-cell analysis (Figure 8) used 10X Genomics droplet-based scRNA-seq, whose lower capture efficiency limits detection of lowly-expressed regulatory factors; the absence of a significant NEO1–HAMP transcriptional correlation at single-cell resolution (ρ = +0.27, P = 0.135) likely reflects both this technical constraint and the inherently protein-level mode of NEO1/BMP/HAMP signaling. Immunohistochemical evidence of NEO1 protein mislocalization in glioma independently supports the conclusion that NEO1 mRNA levels are decoupled from the functional status of this signaling axis. TCGA data provide a static snapshot without direct aspirin exposure information, and gene signatures were used as mechanistic proxies.

## 4. Methods

### 4.1 Study Design and Data Sources

This multi-omics study integrated eight analytical tracks (Figure 6). Transcriptomic data for 172 GBM and 534 LGG samples were obtained from UCSC Xena using RSEM-normalized log2-transformed HiSeqV2 expression values (TCGA GBM/LGG gene expression RNAseq dataset, accessed via UCSC Xena: https://xenabrowser.net/datapages/?dataset=tcga_RSEM_Hugo_norm_count&host=https%3A%2F%2Ftoil.xenahubs.net). CPTAC GBM proteomics reference data were utilized for RNA-protein correlation modeling. TCGA HM450K DNA methylation data were downloaded from UCSC Xena for GBM (n=140) and LGG (n=516). Aspirin perturbation data were derived from GSE198434 and GSE71571. GWAS summary statistics were accessed via IEU OpenGWAS. Single-cell RNA-seq data for 28 GBM patients were obtained from GSE131928 [20].

### 4.2 NEO1/HAMP Axis Analysis

The NEO1/HFE2/BMP6/HAMP signaling axis was characterized by analyzing 36 genes across 7 functional categories: NEO1_Core (NEO1, HFE2, HFE, TFR2, BMP6, TMPRSS6), BMP_SMAD_Signaling (BMPR1A, BMPR2, SMAD1/5/9, SMAD4), HAMP_Regulation (HAMP, SLC40A1, IL6, BMP2), Iron_Uptake (TFRC, STEAP3, SLC11A2, TF), Iron_Storage (FTH1, FTL), Ferroptosis (GPX4, HMOX1, ACSL4, SLC7A11, NFE2L2, AIFM2), and COX_Bridge (PTGS2, PTGES, PTGER2, PTGER4).

### 4.3 DNA Methylation Analysis

TCGA HM450K methylation data for GBM (n=140) and LGG (n=516) were downloaded from UCSC Xena. Probe-to-gene mapping was performed using GPL13534 (HumanMethylation450_15017482_v1.2) platform annotations, successfully mapping 174 probes to promoter regions of NEO1/HAMP axis genes. For genes with multiple probes mapping to the same promoter, the probe with the highest variance across samples was retained to avoid redundant representation. Differential methylation P-values were Bonferroni-corrected for 174 tests. Differential methylation between GBM and LGG was assessed using Mann-Whitney U tests with FDR correction. Methylation-expression concordance was defined as hypomethylation (*Δβ*<0) associated with RNA upregulation or hypermethylation (*Δβ*>0) associated with RNA downregulation.

### 4.4 Gene Set Enrichment Analysis

Preranked GSEA was performed using the signal-to-noise ratio (SNR = [μ_GBM − μ_LGG] / [σ_GBM + σ_LGG]) as the ranking metric across the full transcriptome (≈18,000 genes). Fifteen custom gene sets were tested encompassing NEO1/HAMP signaling, iron metabolism, ferroptosis (driver and suppressor genes), COX/PGE2 pathway, heme metabolism, oxidative stress, BBB tight junctions, inflammatory response, and aspirin-responsive signatures. Enrichment significance was assessed by 1,000 gene-label permutations with NES normalization. FDR was estimated using the Benjamini-Hochberg procedure.

### 4.5 Molecular Subtype Analysis

GBM samples were classified into proneural, mesenchymal, and classical subtypes using the Verhaak classification. LGG samples were stratified by WHO grade (Grade 2 vs. Grade 3) and histology (astrocytoma vs. oligodendroglioma). Expression differences across subtypes were assessed by Kruskal-Wallis test with Dunn’s post-hoc test with Benjamini-Hochberg FDR correction for multiple comparisons.

### 4.6 Additional Methods

COX-2 pathway analysis was performed using differential expression of PTGS2 and its downstream targets (PGE2 synthases PTGES/PTGES2/PTGES3) in GBM versus LGG. Blood-brain barrier (BBB) characterization assessed tight junction protein expression (TJP1/ZO-1, CLDN5, OCLN) across tumor grades. Aspirin-response gene signature projection used published gene sets (COX2_Glioma_Prog, Aspirin_GBM_5-gene) to stratify glioma survival via ssGSEA. BBB integrity evaluation and ferroptosis vulnerability scoring (mean expression of 15 driver genes minus 20 suppressor genes) were computed as described in Section 2.8. Mendelian randomization (MR) analysis was performed using the TwoSampleMR R package (v0.5.6) [13] with five PTGS2 cis-SNPs as instrumental variables (F-statistic > 10), applying inverse-variance weighted (IVW) random-effects meta-analysis as primary method, with MR-Egger, weighted median, and Cochran Q heterogeneity test as sensitivity analyses. All statistical analyses were performed in R (v4.2.0) and Python (v3.10) using two-sided tests with P < 0.05 considered significant unless otherwise specified. The multi-gene COX risk score (PTGS2 + PTGES + PTGER2 + PTGER4) was calculated as a weighted linear combination with weights derived from univariate Cox regression coefficients.

### 4.7 Single-Cell RNA-seq Validation

Single-cell RNA-seq data from 28 GBM patients (GSE131928, Neftel et al. [20]) were obtained from the Gene Expression Omnibus. Processed TPM expression matrices were downloaded from GEO (GSE131928). We restricted analysis to the 10X Genomics platform data (16,201 cells) to ensure uniform capture efficiency and data distribution. Cells were filtered to retain those with >200 expressed genes, >500 UMIs, and <20% mitochondrial reads, retaining 15,112 high-quality malignant cells for downstream analysis. Normalization was performed to 10,000 transcripts per cell followed by log-transformation. Highly variable genes (n=4,000) were identified using the Seurat v3 flavor [21]. Principal component analysis (50 components) was performed on scaled data, and the first 30 principal components were used for UMAP dimensionality reduction [22] and neighborhood construction (k=20). Malignant cell state classification was performed using the Neftel 2019 meta-module gene signatures [20] with sc.tl.score_genes(), assigning each cell to the state (MES, AC, OPC, or NPC) with the highest module score. Ferroptosis vulnerability scores were calculated as the difference between the mean expression of driver genes (ACSL4, HMOX1, PTGS2, LPCAT3, ALOX5, ALOX12, ALOX15, POR, CYB5R1, NOX1) and suppressor genes (GPX4, SLC7A11, NFE2L2, FTH1, FTL, SLC3A2, GCH1, DHODH, AIFM2, FSP1, COQ2, COQ5, HSPB1). Differences across cell states were assessed by Mann-Whitney U tests. Single-cell expression correlations were evaluated using Spearman rank correlation coefficient.

### 4.8 Virtual Drug Screening

Drug virtual screening integrated three complementary approaches. First, a drug-target interaction matrix was curated from DGIdb v4.0 (https://dgidb.org), ferroptosis pharmacology literature, and published drug-HAMP regulatory data, covering 35 interactions across 21 unique drugs targeting 9 signature genes. Drug priority scores were calculated as a weighted composite of: (i) number of signature genes targeted; (ii) directional concordance (upregulated genes targeted by inhibitors, downregulated genes targeted by activators); (iii) interaction-level evidence weighting (direct molecular target = 5×, pathway-level effect = 2×, literature-reported = 1×); and (iv) pharmacological category bonus (ferroptosis inducer = 3×, aspirin pathway = 3×, HAMP modulator = 2×, iron chelator = 1×). Second, GDSC2 drug sensitivity data (https://www.cancerrxgene.org) were queried for 7 glioma cell lines (U87MG, U251MG, T98G, LN229, A172, U118MG, SF268) to identify ferroptosis-inducing agents with selective activity against iron-high GBM models. Third, the drug-gene interaction matrix was visualized as a multi-panel figure comprising: priority score bar chart (top 20 agents), drug-gene target matrix, aspirin mechanistic bridge schematic, and ferroptosis inducer-target interaction heatmap (Figure 9).

## 5. Conclusions

This multi-omics integrative analysis provides convergent evidence across transcriptomic, epigenomic, pathway-level enrichment, single-cell transcriptomic, drug screening, and causal inference domains supporting a central role for the NEO1/hepcidin iron regulatory axis in glioma (Figure 7). De novo HM450K analysis identified HAMP as the dominant epigenetically regulated target (Δβ=-0.265; Bonferroni-corrected P=2.5×10⁻⁴⁶) and revealed a baseline hypomethylation state for NEO1, TFRC, and GPX4 in brain tissue. GSEA confirmed ferroptosis driver genes as the most significantly enriched pathway, and molecular subtype analysis identified the mesenchymal GBM subtype as the iron-high phenotype. Single-cell RNA-seq analysis across 28 GBM patients validated the heterogeneous distribution of iron metabolism gene expression across malignant cell states and independently confirmed the mesenchymal state as the iron-high, ferroptosis-vulnerable compartment. Universal GPX4 overexpression across all cell states identifies this ferroptosis suppressor as a pan-GBM dependency with therapeutic relevance. Multi-pathway PCD analysis revealed coordinated ferroptosis-PANoptosis activation with reciprocal pyroptosis suppression in GBM, extending the cell death vulnerability framework beyond ferroptosis alone. Visium spatial transcriptomic validation independently confirmed the NEO1 edge-high gradient, GPX4 pan-tumor expression, and TFRC core-high pattern at the tissue architecture level. PTGS2 serves as a molecular bridge connecting COX-2 inhibition, ferroptosis vulnerability, and aspirin’s chemopreventive effects. Virtual drug screening identified 14 of 17 NEO1/HAMP axis genes as druggable by existing agents, with GPX4 inhibitors (RSL3, FIN56) emerging as the highest-priority candidates for targeting the pan-GBM ferroptosis dependency we identified at single-cell resolution. These findings establish a comprehensive multi-omics-to-therapeutics framework for future clinical and mechanistic investigation of iron metabolism-targeted therapies in neuro-oncology.

## List of Abbreviations

BBB: blood-brain barrier; BMP: bone morphogenetic protein; CGGA: Chinese Glioma Genome Atlas; COX: cyclooxygenase; CPTAC: Clinical Proteomic Tumor Analysis Consortium; FDR: false discovery rate; GBM: glioblastoma; GEO: Gene Expression Omnibus; GPX4: glutathione peroxidase 4; GSEA: gene set enrichment analysis; GWAS: genome-wide association study; HAMP: hepcidin antimicrobial peptide; HFE: homeostatic iron regulator; HFE2: hemojuvelin; HM450K: HumanMethylation450 BeadChip; HMOX1: heme oxygenase 1; HR: hazard ratio; IVW: inverse-variance weighted; LGG: lower-grade glioma; MES: mesenchymal; MR: Mendelian randomization; NEO1: neogenin 1; NES: normalized enrichment score; PCD: programmed cell death; PGE2: prostaglandin E2; PTGS2: prostaglandin-endoperoxide synthase 2 (COX-2); scRNA-seq: single-cell RNA sequencing; ssGSEA: single-sample gene set enrichment analysis; TCGA: The Cancer Genome Atlas; TF: transcription factor; TFRC: transferrin receptor

## Declarations

### Ethics approval and consent to participate

This study is a retrospective analysis of publicly available de-identified datasets from TCGA, CPTAC, GEO, and IEU OpenGWAS. No new human data, patient samples, or clinical interventions were involved. Ethics approval and informed consent were obtained by the original data-generating consortia. Therefore, additional ethics approval is not applicable to this study.

### Consent for publication

Consent for publication was obtained by the original data-generating consortia. No individual person’s data are presented in this study.

### Availability of data and materials

All data used in this study are publicly available. TCGA transcriptomic and methylation data were accessed via UCSC Xena (https://xenabrowser.net). CPTAC proteomics data are available via the CPTAC portal (https://cptac-data-portal.georgetown.edu). GEO datasets (GSE131928, GSE198434, GSE71571) were accessed from the NCBI Gene Expression Omnibus (https://www.ncbi.nlm.nih.gov/geo). GWAS summary statistics were obtained from IEU OpenGWAS (https://gwas.mrcieu.ac.uk). All analysis scripts are available from the corresponding author upon reasonable request.

### Competing interests

The authors declare that they have no competing interests.

### Funding

This research did not receive any specific grant from funding agencies in the public, commercial, or not-for-profit sectors.

### Authors’ contributions

Conceptualization, X.W.; Methodology, X.W., C.M., F.Z.; Software, F.W., C.S.; Formal Analysis, C.M., F.Z., F.W.; Investigation, X.W., C.S., X.T.; Data Curation, C.M., F.Z.; Writing — Original Draft, X.W.; Writing — Review & Editing, all authors; Supervision, X.W.; Project Administration, X.W. All authors have read and approved the final manuscript.

## Data Availability

All data used in this study are publicly available. TCGA transcriptomic and methylation data were accessed via UCSC Xena (https://xenabrowser.net). CPTAC proteomics data are available via the CPTAC portal (https://cptac-data-portal.georgetown.edu). GEO datasets were accessed from the NCBI Gene Expression Omnibus (https://www.ncbi.nlm.nih.gov/geo). GWAS summary statistics were obtained from IEU OpenGWAS (https://gwas.mrcieu.ac.uk). Single-cell RNA-seq data (GSE131928) are available from GEO. All analysis scripts are provided in the project repository.

## Acknowledgements

Not applicable.

## Supplementary Materials

Figure S1. Additional Visium spatial expression maps for HAMP, PTGS2, and SLC7A11 in human glioblastoma (see Section 2.12).

Figure S2. TCGA microbiome-iron metabolism axis analysis in GBM (composite panel). (A) Top 10 most prevalent bacterial families in GBM tumors. (B) Heatmap of Spearman correlations between bacterial family abundance and iron metabolism gene expression. (C) Scatter plots of key bacterial genera vs. NEO1, HAMP, and TFRC expression.

Figure S3. Transcription factor regulatory network and virtual NEO1 knockout simulation. (A) Spearman correlation heatmap of 41 transcription factors vs. 8 iron metabolism genes in TCGA GBM (n=172). (B) Virtual NEO1 knockout simulation: predicted impact on the 25-gene iron regulatory network based on co-expression GRN.

Figure S4. Independent CGGA external validation and xCell immune deconvolution. (A) CGGA expression direction concordance (13/13 genes, 100%). (B-C) xCell immune cell type correlation heatmaps for iron metabolism genes in TCGA (B) and CGGA (C). (D) CGGA Cox regression forest plot.

Figure S5. Machine learning integration and IDH stratification (composite panel). (A) LASSO Cox coefficient path. (B) Final LASSO Cox coefficients showing NEO1 as protective (negative coefficient), STEAP3/TFRC/HFE as risk genes (C-index=0.827). (C) Random Forest feature importance ranking (CV AUC=0.947). (D) RF ROC curve.

**Supplementary Table S1.** Gene Set Enrichment Analysis results - all 15 gene sets with NES, P-value, and FDR (provided in supplementary_materials.docx).

**Supplementary Table S2.** Multi-omics evidence matrix with integrated scoring (provided in supplementary_materials.docx).

**Supplementary Table S3.** Complete Spearman correlation results between bacterial family abundance and iron metabolism gene expression (n=161 GBM patients).

**Supplementary Table S4.** Transcription factor-target gene correlation matrix (41 TFs x 8 targets) and gene regulatory network edge list (60 nodes).

**Supplementary Table S5.** CGGA univariate Cox regression results (n=693 glioma samples).

**Supplementary Table S6.** xCell immune deconvolution scores and gene-immune correlations for TCGA and CGGA cohorts.

## Notes

### Competing Interest Statement

The authors have declared no competing interest.

### Author Declarations

The study used ONLY openly available human data. All data sources were publicly accessible before the initiation of the study: TCGA data via UCSC Xena (https://xenabrowser.net), CPTAC data via the CPTAC portal (https://cptac-data-portal.georgetown.edu), GEO datasets (https://www.ncbi.nlm.nih.gov/geo), GWAS data from IEU OpenGWAS (https://gwas.mrcieu.ac.uk), and single-cell RNA-seq data from GEO (GSE131928).

### Summary of Updates

Data Consistency and Sample Size Clarification: We explicitly clarified the discrepancy in PCD analysis sample sizes (702 vs 706 total TCGA samples). We now specify that 4 LGG samples were excluded after rigorous quality control filtering (530 LGG retained from 534 total), ensuring alignment between section 2.13 and the abstract. Epigenetic Statistical Rigor: In response to requests for stricter statistical validation, we supplemented the HAMP methylation analysis (delta beta = -0.265, P = 1.4 x 10^-48) with Bonferroni-corrected P-values and FDR q-values. We confirm that HAMP remains the dominant epigenetic target after multiple testing correction. Single-Cell Methodology Transparency: We resolved inconsistencies regarding GSE131928 single-cell data. We explicitly state that our analysis focused exclusively on 10X Genomics data (15,112 cells) following QC filtering of Smart-seq2 data, aligning with the reported cell count. We expanded the discussion to interpret the non-significant NEO1-HAMP correlation (Spearman rho = +0.27, P = 0.135) in the context of protein-level signaling complexity and technical limitations of droplet-based scRNA-seq. Reference Verification and Standardization: All references cited in the discussion, including Chen et al. (Nature 2026) and Dong et al. (bioRxiv 2026), have been verified and updated with complete DOIs, PMIDs, and standardized citation formats. Preprint citations are clearly labeled as non-peer-reviewed. Methodological Detail Expansion: We detailed the multi-gene COX risk score calculation (weighted by univariate Cox coefficients) and clarified the RNA-protein concordance analysis in CPTAC as a correlation modeling approach rather than direct quantitative proteomics. Mechanistic Interpretation Refinement: We refined interpretations regarding NEO1/HAMP axis regulation, emphasizing that while transcriptional and epigenetic dysregulation occurs, protein mislocalization (supported by immunohistochemistry in companion work) decouples mRNA levels from functional signaling output.

