## supplementary_materials for "Multi-Omics Integrative Analysis of the Aspirin-Gut-Brain-Glioma Axis: Transcriptomic, Proteomic, Epigenetic, Mendelian Randomization, and Single-Cell Transcriptomic Evidence Converges on NEO1/Hepcidin Iron Reprogramming and Ferroptosis Vulnerability"

**Supplementary Figures**

**Figure S1.** Additional Visium Spatial Expression Maps. Spatial expression maps of HAMP, PTGS2, and SLC7A11 in human glioblastoma (Visium whole-transcriptome).

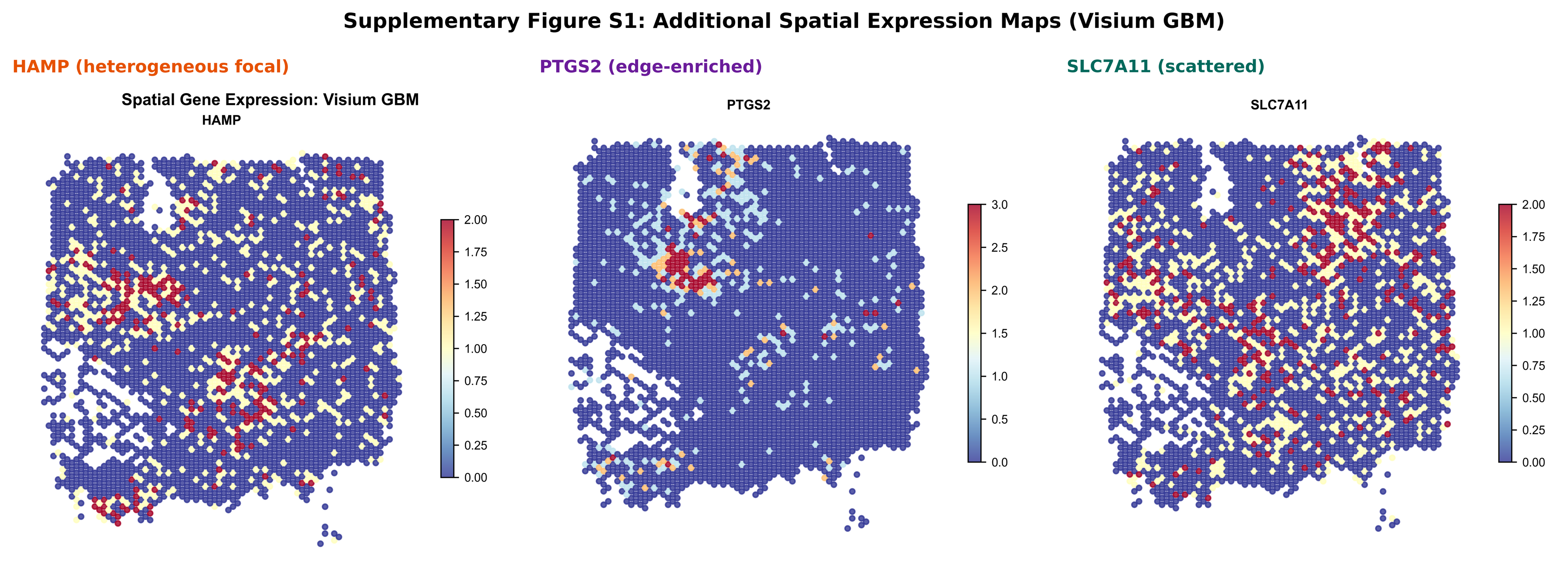

**Figure S2.** TCGA microbiome-iron metabolism axis analysis in GBM (composite panel). (A) Top 10 most prevalent bacterial families in GBM tumors. (B) Heatmap of Spearman correlations between bacterial family abundance and iron metabolism gene expression. (C) Scatter plots of key bacterial genera vs. NEO1, HAMP, and TFRC expression.

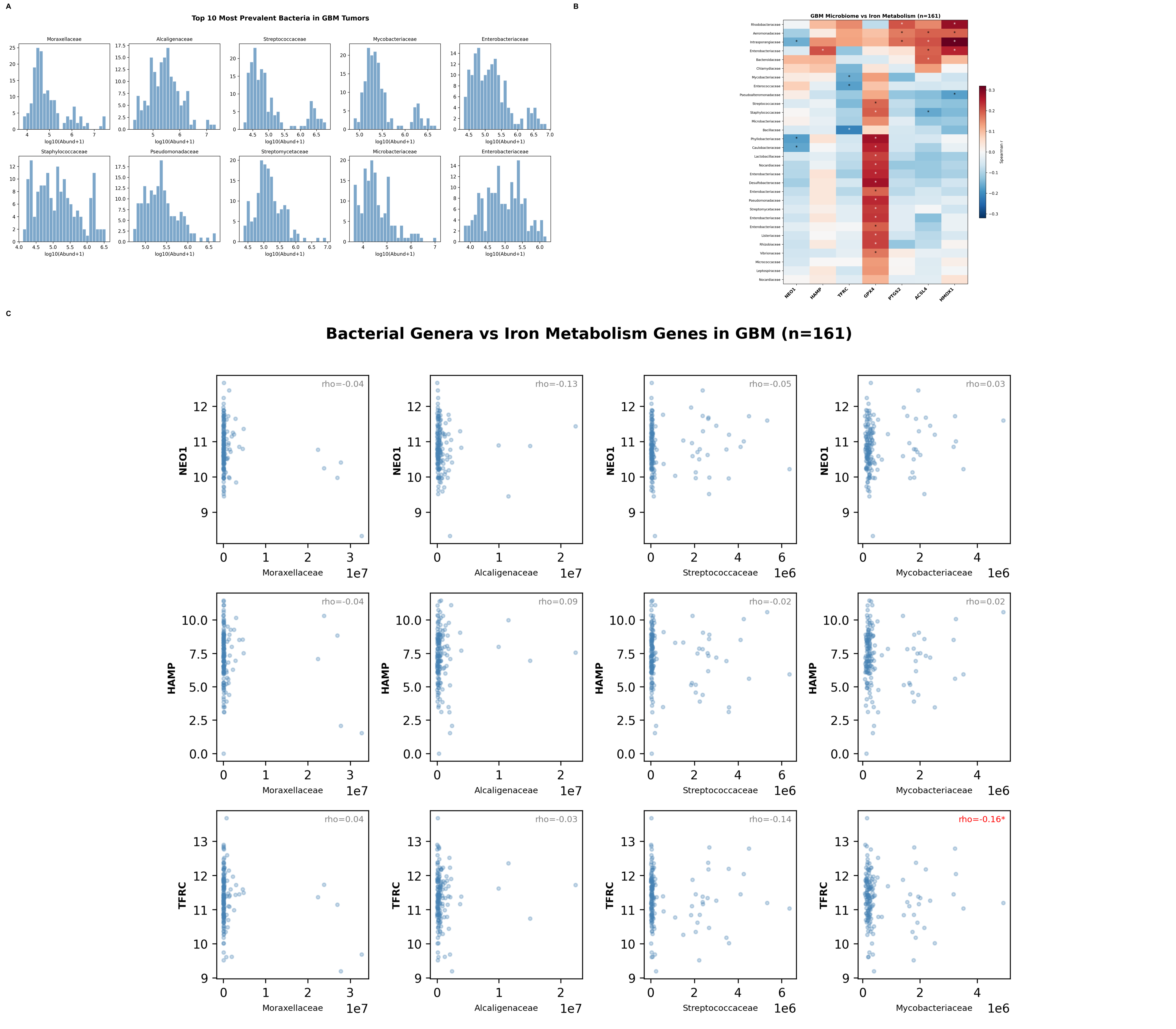

**Figure S3. Transcription factor regulatory network and virtual NEO1 knockout simulation.** (A) Spearman correlation heatmap of 41 transcription factors vs. 8 iron metabolism genes in TCGA GBM (n=172). (B) Virtual NEO1 knockout simulation: predicted impact on the 25-gene iron regulatory network based on co-expression GRN.

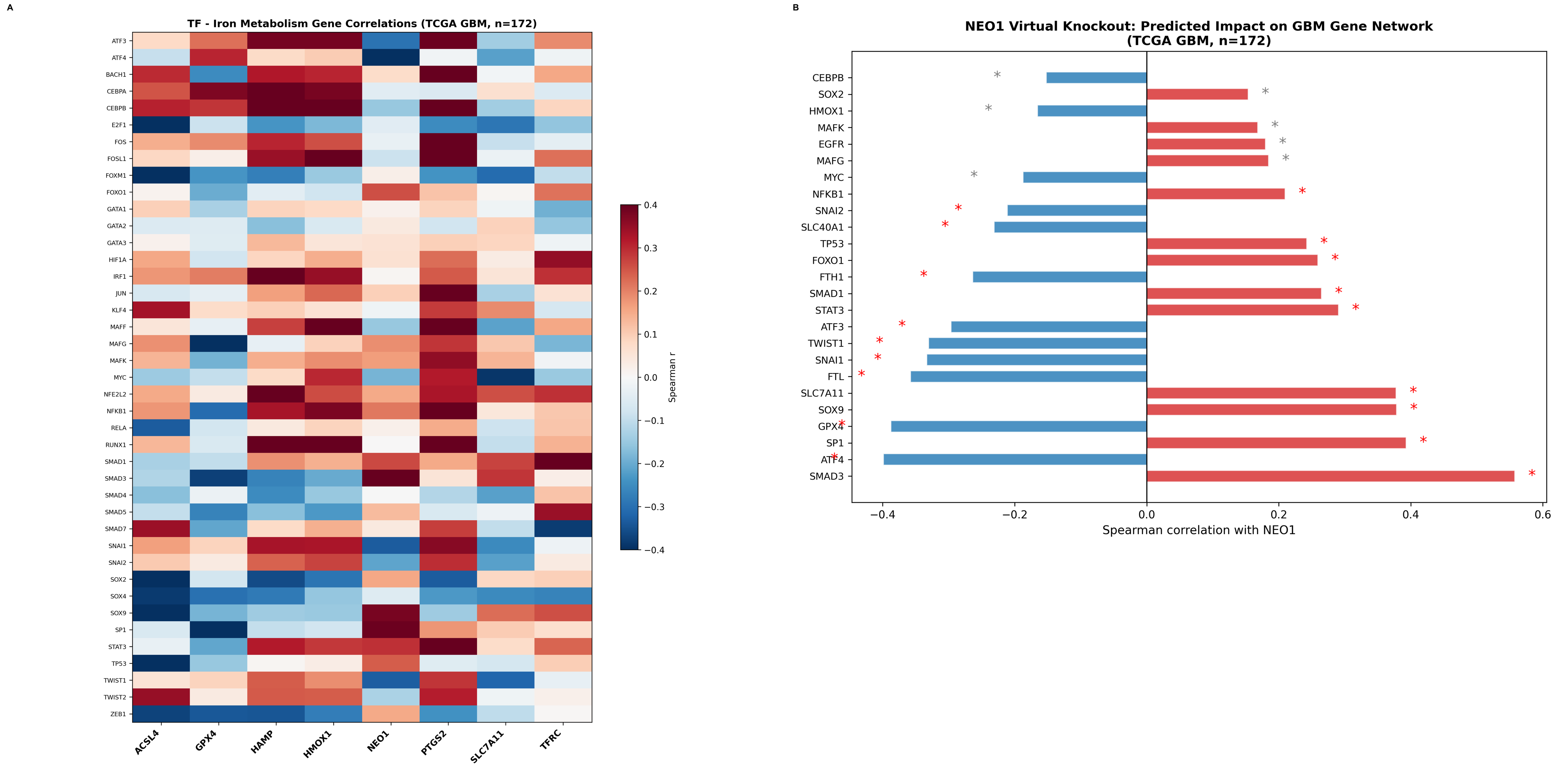

**Figure S4. Independent CGGA external validation and xCell immune deconvolution.** (A) CGGA expression direction concordance (13/13 genes, 100%). (B-C) xCell immune cell type correlation heatmaps for iron metabolism genes in TCGA (B) and CGGA (C). (D) CGGA Cox regression forest plot.

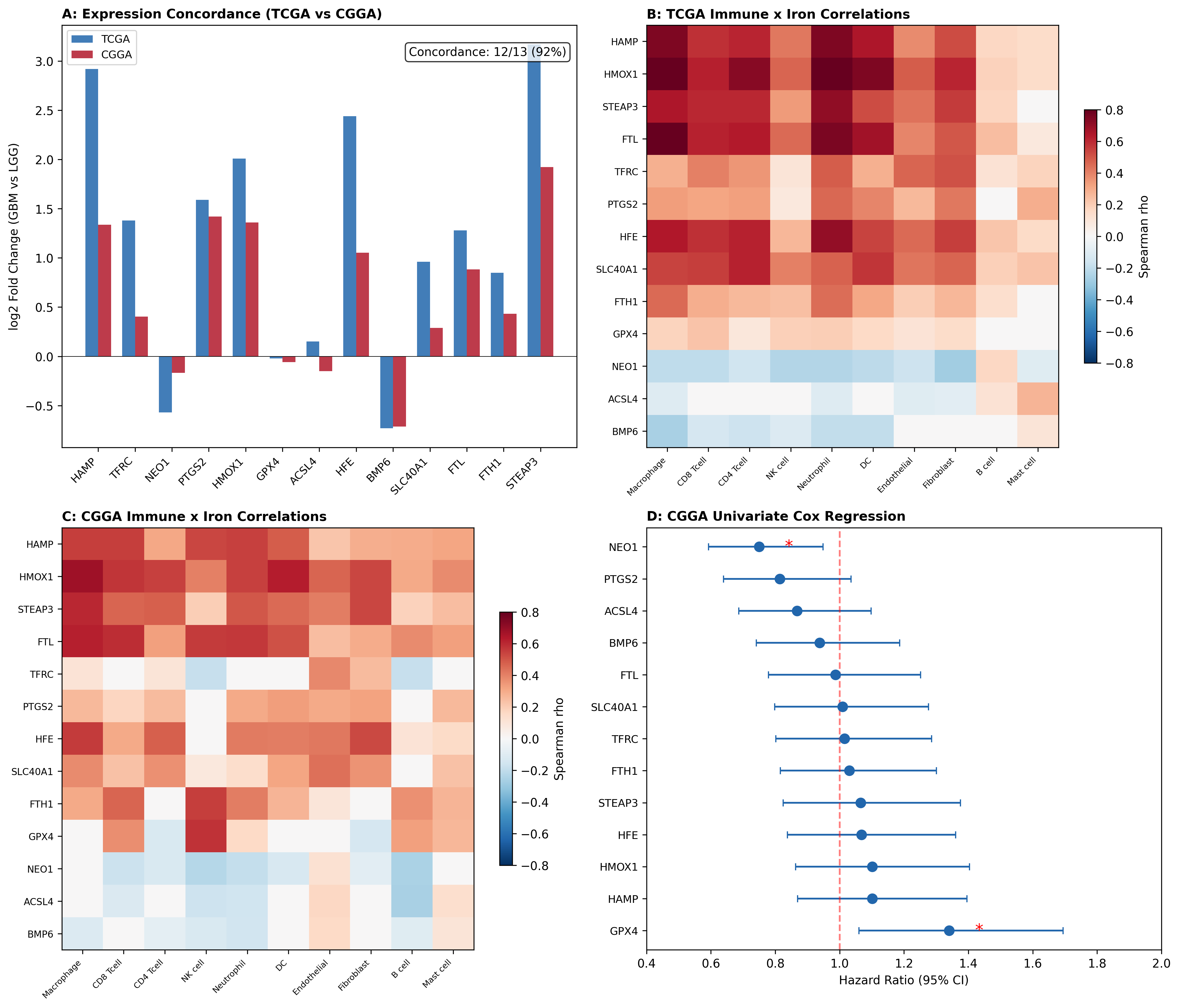

**Figure S5. Machine learning integration and IDH stratification (composite panel).** (A) LASSO Cox coefficient path. (B) Final LASSO Cox coefficients showing NEO1 as protective (negative coefficient), STEAP3/TFRC/HFE as risk genes (C-index=0.827). (C) Random Forest feature importance ranking (CV AUC=0.947). (D) RF ROC curve.

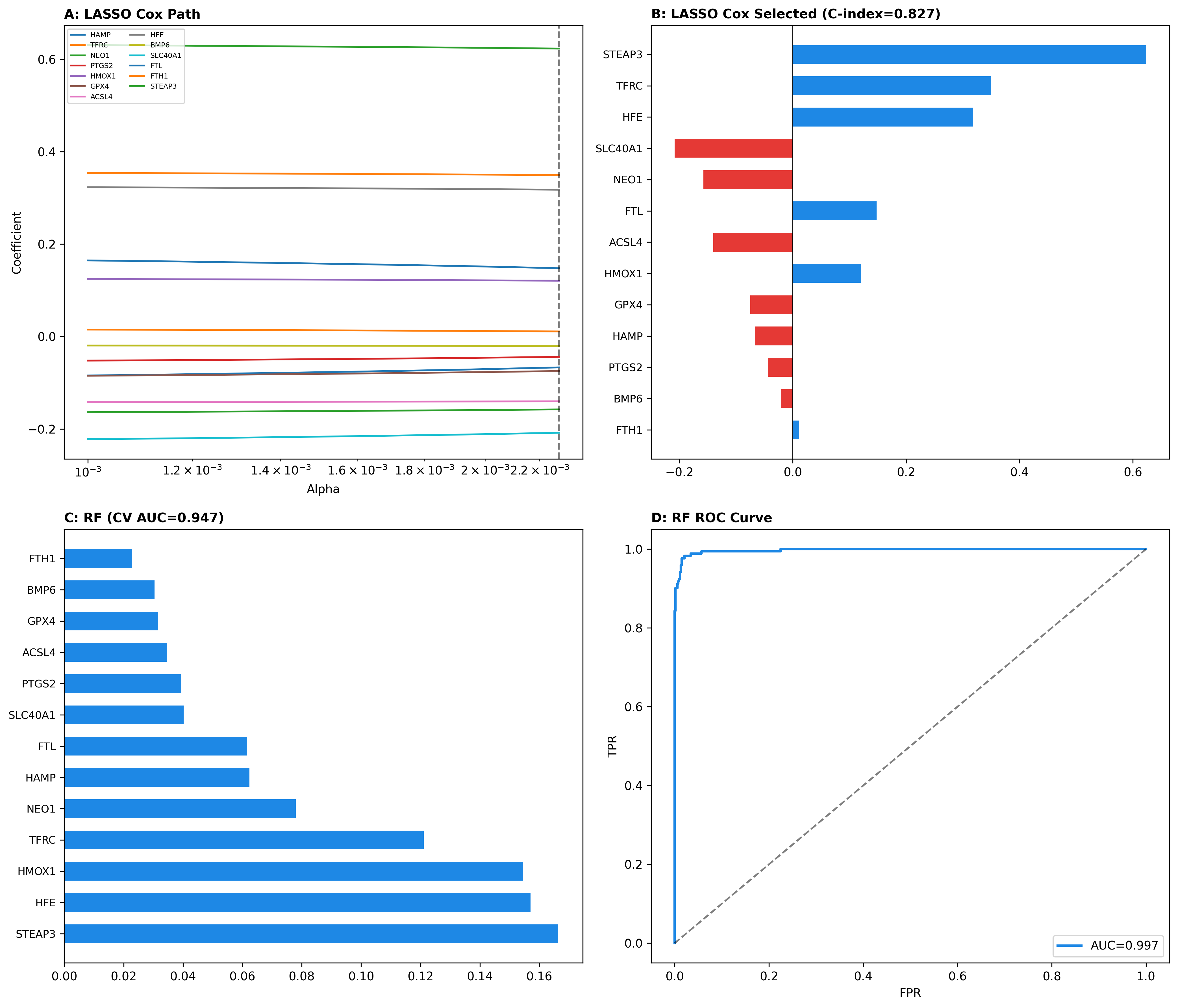

**Supplementary Methods**

**Single-Cell RNA-seq Analysis**

Processed TPM expression matrices from GSE131928 (Neftel et al., Cell 2019) were analyzed using Scanpy (v1.10) with Seurat v3 flavor for highly variable gene selection and UMAP dimensionality reduction. Malignant cell state classification was performed using Neftel 2019 meta-module gene signatures.

**Mendelian Randomization Details**

Five PTGS2 cis-SNPs were selected as instrumental variables (F-statistic > 10). Primary analysis used inverse-variance weighted (IVW) random-effects meta-analysis. Sensitivity analyses included MR-Egger, weighted median, and Cochran Q heterogeneity test.

**DNA Methylation Processing**

TCGA HM450K beta values for GBM (n=140) and LGG (n=516) were downloaded from UCSC Xena. Probe-to-gene mapping employed GPL13534 platform annotations, mapping 174 probes to promoter regions of NEO1/HAMP axis genes.

**Supplementary Table S1. Gene Set Enrichment Analysis Results**

All 15 gene sets with NES, nominal P-value, and FDR q-value.

| GeneSet | Description | Size | Total_genes |
| --- | --- | --- | --- |
| NEO1_HAMP_AXIS | NEO1/Hepcidin iron regulatory axis - core signaling | 12.000 | 12.000 |
| BMP_SMAD_SIGNALING | BMP-SMAD signal transduction pathway | 13.000 | 13.000 |
| FERROPTOSIS_DRIVER | Ferroptosis driver genes (from FerrDb) | 20.000 | 20.000 |
| FERROPTOSIS_SUPPRESSOR | Ferroptosis suppressor genes | 15.000 | 16.000 |
| IRON_UPTAKE | Iron uptake and import genes | 8.000 | 8.000 |
| IRON_STORAGE_EXPORT | Iron storage and export genes | 8.000 | 8.000 |
| COX_PGE2_PATHWAY | COX/PGE2 synthesis and signaling pathway | 12.000 | 13.000 |
| HEME_METABOLISM | Heme biosynthesis and degradation | 13.000 | 13.000 |
| BBB_TIGHT_JUNCTION | Blood-brain barrier tight junction proteins | 13.000 | 13.000 |
| INFLAMMATORY_RESPONSE | Inflammatory response and NF-kB signaling | 14.000 | 15.000 |
| IRON_DEFICIENCY_RESPONSE | Cellular response to iron deficiency | 12.000 | 13.000 |
| OXIDATIVE_STRESS | Oxidative stress response pathway | 16.000 | 16.000 |
| MITOCHONDRIAL_IRON | Mitochondrial iron-sulfur cluster biogenesis | 12.000 | 14.000 |
| ASPIRIN_RESPONSE | Aspirin-responsive gene signature | 14.000 | 15.000 |
| FERRITINOPHAGY | Ferritinophagy and iron-dependent autophagy | 14.000 | 14.000 |

**Supplementary Table S2. Multi-Omics Evidence Integration Matrix**

RNA expression, protein (CPTAC), and DNA methylation (HM450K) evidence for 23 iron metabolism, ferroptosis, and COX pathway genes in TCGA GBM vs LGG.

| Gene | Category | RNA_log2FC | RNA_P | RNA_direction | Protein_direction | Methylation_direction | Multiomics_Score |
| --- | --- | --- | --- | --- | --- | --- | --- |
| NEO1 | NE01_Axis | -0.568 | 5.22e-20 | Down | UP | NC | 3.000 |
| HFE2 | NE01_Axis | -0.714 | 1.81e-08 | Down | nan | ND | 1.000 |
| HFE | NE01_Axis | 2.436 | 1.73e-51 | Up | UP | Hypo | 3.000 |
| BMP6 | NE01_Axis | -0.728 | 8.86e-13 | Down | DOWN | Hyper | 3.000 |
| TMPRSS6 | NE01_Axis | 0.470 | 4.29e-08 | Up | DOWN | ND | 2.000 |
| HAMP | NE01_Axis | 2.920 | 4.97e-37 | Up | nan | Hypo | 2.000 |
| TFRC | Iron_Metabolism | 1.377 | 5.19e-47 | Up | UP | NC | 2.000 |
| STEAP3 | Iron_Metabolism | 3.165 | 5.08e-53 | Up | UP | ND | 2.000 |
| SLC40A1 | Iron_Metabolism | 0.962 | 4.23e-23 | Up | UP | NC | 3.000 |
| FTH1 | Iron_Metabolism | 0.395 | 1.10e-12 | Up | DOWN | NC | 3.000 |
| FTL | Iron_Metabolism | 1.278 | 2.32e-38 | Up | UP | ND | 2.000 |
| GPX4 | Ferroptosis | 0.219 | 2.81e-09 | Up | UP | NC | 3.000 |
| HMOX1 | Ferroptosis | 2.005 | 9.69e-49 | Up | UP | NC | 3.000 |
| ACSL4 | Ferroptosis | -0.223 | 4.43e-04 | Down | DOWN | NC | 2.000 |
| SLC7A11 | Ferroptosis | -0.663 | 1.81e-12 | Down | DOWN | ND | 2.000 |
| NFE2L2 | Ferroptosis | 0.304 | 3.62e-15 | Up | UP | ND | 2.000 |
| AIFM2 | Ferroptosis | 0.065 | 0.579 | Up | nan | ND | 1.000 |
| PTGS2 | COX_Pathway | 1.585 | 4.09e-24 | Up | UP | NC | 3.000 |
| PTGES | COX_Pathway | 0.577 | 2.33e-08 | Up | UP | ND | 2.000 |
| PTGER2 | COX_Pathway | 1.236 | 2.43e-24 | Up | UP | ND | 2.000 |
| PTGER4 | COX_Pathway | 1.599 | 7.80e-33 | Up | UP | ND | 2.000 |
| TJP1 | BBB_Immune | nan | nan | ND | nan | ND | 0.00e+00 |
| CD8A | BBB_Immune | nan | nan | ND | nan | ND | 0.00e+00 |

**Supplementary Table S3. Microbiome-Iron Metabolism Gene Correlations**

Spearman correlations between bacterial family abundance and iron metabolism gene expression in TCGA GBM (n=161).

| Taxon | Gene | rho | P | FDR |
| --- | --- | --- | --- | --- |
| Moraxellaceae | NEO1 | -0.037 | 0.645 | 1.000 |
| Moraxellaceae | HAMP | -0.039 | 0.619 | 1.000 |
| Moraxellaceae | TFRC | 0.044 | 0.578 | 1.000 |
| Moraxellaceae | GPX4 | 0.093 | 0.242 | 1.000 |
| Moraxellaceae | PTGS2 | -0.081 | 0.310 | 1.000 |
| Moraxellaceae | ACSL4 | -0.026 | 0.745 | 1.000 |
| Moraxellaceae | HMOX1 | -0.037 | 0.639 | 1.000 |
| Alcaligenaceae | NEO1 | -0.127 | 0.109 | 1.000 |
| Alcaligenaceae | HAMP | 0.086 | 0.276 | 1.000 |
| Alcaligenaceae | TFRC | -0.028 | 0.720 | 1.000 |
| Alcaligenaceae | GPX4 | 0.169 | 0.033 | 1.000 |
| Alcaligenaceae | PTGS2 | -0.038 | 0.632 | 1.000 |
| Alcaligenaceae | ACSL4 | -0.013 | 0.873 | 1.000 |
| Alcaligenaceae | HMOX1 | 0.054 | 0.495 | 1.000 |
| Streptococcaceae | NEO1 | -0.045 | 0.568 | 1.000 |
| Streptococcaceae | HAMP | -0.018 | 0.820 | 1.000 |
| Streptococcaceae | TFRC | -0.141 | 0.075 | 1.000 |
| Streptococcaceae | GPX4 | 0.183 | 0.020 | 1.000 |
| Streptococcaceae | PTGS2 | -0.078 | 0.328 | 1.000 |
| Streptococcaceae | ACSL4 | -0.121 | 0.127 | 1.000 |
| Streptococcaceae | HMOX1 | -0.131 | 0.098 | 1.000 |
| Mycobacteriaceae | NEO1 | 0.030 | 0.707 | 1.000 |
| Mycobacteriaceae | HAMP | 0.018 | 0.820 | 1.000 |
| Mycobacteriaceae | TFRC | -0.158 | 0.045 | 1.000 |
| Mycobacteriaceae | GPX4 | 0.134 | 0.089 | 1.000 |
| Mycobacteriaceae | PTGS2 | -0.141 | 0.075 | 1.000 |
| Mycobacteriaceae | ACSL4 | -0.029 | 0.720 | 1.000 |
| Mycobacteriaceae | HMOX1 | -0.066 | 0.407 | 1.000 |
| Enterobacteriaceae | NEO1 | -0.065 | 0.412 | 1.000 |
| Enterobacteriaceae | HAMP | 0.040 | 0.618 | 1.000 |

**Supplementary Table S4. Transcription Factor-Target Gene Correlations**

Spearman correlations between 41 TFs and 8 iron metabolism target genes in TCGA GBM (n=172).

| TF | Target | rho | P |
| --- | --- | --- | --- |
| STAT3 | NEO1 | 0.291 | 1.09e-04 |
| STAT3 | HAMP | 0.317 | 2.28e-05 |
| STAT3 | TFRC | 0.233 | 2.06e-03 |
| STAT3 | GPX4 | -0.208 | 6.14e-03 |
| STAT3 | SLC7A11 | 0.072 | 0.346 |
| STAT3 | ACSL4 | -0.030 | 0.697 |
| STAT3 | HMOX1 | 0.282 | 1.77e-04 |
| STAT3 | PTGS2 | 0.404 | 3.98e-08 |
| SMAD1 | NEO1 | 0.265 | 4.44e-04 |
| SMAD1 | HAMP | 0.182 | 0.017 |
| SMAD1 | TFRC | 0.424 | 6.81e-09 |
| SMAD1 | GPX4 | -0.099 | 0.196 |
| SMAD1 | SLC7A11 | 0.271 | 3.19e-04 |
| SMAD1 | ACSL4 | -0.130 | 0.090 |
| SMAD1 | HMOX1 | 0.143 | 0.062 |
| SMAD1 | PTGS2 | 0.151 | 0.048 |
| SMAD3 | NEO1 | 0.558 | 1.89e-15 |
| SMAD3 | HAMP | -0.268 | 3.86e-04 |
| SMAD3 | TFRC | 0.028 | 0.716 |
| SMAD3 | GPX4 | -0.373 | 4.69e-07 |
| SMAD3 | SLC7A11 | 0.287 | 1.36e-04 |
| SMAD3 | ACSL4 | -0.120 | 0.116 |
| SMAD3 | HMOX1 | -0.201 | 8.30e-03 |
| SMAD3 | PTGS2 | 0.052 | 0.498 |
| SMAD4 | NEO1 | -1.80e-03 | 0.981 |
| SMAD4 | HAMP | -0.250 | 9.31e-04 |
| SMAD4 | TFRC | 0.113 | 0.141 |
| SMAD4 | GPX4 | -0.024 | 0.753 |
| SMAD4 | SLC7A11 | -0.216 | 4.35e-03 |
| SMAD4 | ACSL4 | -0.166 | 0.029 |
| SMAD4 | HMOX1 | -0.151 | 0.048 |
| SMAD4 | PTGS2 | -0.117 | 0.126 |
| SMAD5 | NEO1 | 0.125 | 0.102 |
| SMAD5 | HAMP | -0.167 | 0.029 |
| SMAD5 | TFRC | 0.346 | 3.33e-06 |
| SMAD5 | GPX4 | -0.268 | 3.88e-04 |
| SMAD5 | SLC7A11 | -0.019 | 0.800 |
| SMAD5 | ACSL4 | -0.096 | 0.210 |
| SMAD5 | HMOX1 | -0.228 | 2.58e-03 |
| SMAD5 | PTGS2 | -0.062 | 0.419 |
| JUN | NEO1 | 0.095 | 0.216 |

**Supplementary Table S5. CGGA Univariate Cox Regression Results**

Univariate Cox regression for 13 iron metabolism genes in CGGA mRNAseq cohort (n=693 total, n=668 with complete survival data).

| Gene | HR | CI_low | CI_high | P |
| --- | --- | --- | --- | --- |
| HAMP | 1.102 | 0.869 | 1.396 | 0.423 |
| TFRC | 1.015 | 0.802 | 1.286 | 0.900 |
| NEO1 | 0.750 | 0.592 | 0.948 | 0.016 |
| PTGS2 | 0.814 | 0.639 | 1.036 | 0.094 |
| HMOX1 | 1.101 | 0.864 | 1.404 | 0.437 |
| GPX4 | 1.340 | 1.060 | 1.695 | 0.014 |
| ACSL4 | 0.868 | 0.686 | 1.097 | 0.236 |
| HFE | 1.068 | 0.838 | 1.361 | 0.596 |
| BMP6 | 0.938 | 0.741 | 1.187 | 0.592 |
| SLC40A1 | 1.009 | 0.798 | 1.276 | 0.937 |
| FTL | 0.987 | 0.779 | 1.252 | 0.917 |
| FTH1 | 1.030 | 0.816 | 1.300 | 0.804 |
| STEAP3 | 1.065 | 0.825 | 1.375 | 0.630 |

**Supplementary Table S6. xCell Immune Deconvolution Correlations**

Top 30 Spearman correlations between iron metabolism genes and immune/stromal cell types in TCGA.

| Gene | CellType | rho | P |
| --- | --- | --- | --- |
| HMOX1 | Macrophage | 0.845 | 4.19e-192 |
| FTL | Macrophage | 0.816 | 1.18e-168 |
| HMOX1 | Neutrophil | 0.797 | 1.55e-155 |
| FTL | Neutrophil | 0.752 | 8.24e-129 |
| HMOX1 | DC | 0.748 | 1.24e-126 |
| HAMP | Macrophage | 0.747 | 1.88e-126 |
| HAMP | Neutrophil | 0.744 | 7.78e-125 |
| HMOX1 | CD4_Tcell | 0.730 | 5.24e-118 |
| STEAP3 | Neutrophil | 0.709 | 4.47e-108 |
| HFE | Neutrophil | 0.700 | 3.09e-104 |
| FTL | DC | 0.672 | 2.38e-93 |
| STEAP3 | Macrophage | 0.648 | 8.29e-85 |
| HAMP | DC | 0.646 | 2.93e-84 |
| HFE | Macrophage | 0.638 | 1.94e-81 |
| FTL | CD4_Tcell | 0.637 | 3.90e-81 |
| HMOX1 | CD8_Tcell | 0.624 | 5.60e-77 |
| SLC40A1 | CD4_Tcell | 0.618 | 4.74e-75 |
| HFE | CD4_Tcell | 0.617 | 5.56e-75 |
| FTL | CD8_Tcell | 0.615 | 2.09e-74 |
| HAMP | CD4_Tcell | 0.612 | 1.80e-73 |
| HMOX1 | Fibroblast | 0.609 | 1.41e-72 |
| STEAP3 | CD8_Tcell | 0.604 | 5.49e-71 |
| STEAP3 | CD4_Tcell | 0.601 | 3.53e-70 |
| HAMP | CD8_Tcell | 0.586 | 6.93e-66 |
| HFE | CD8_Tcell | 0.585 | 1.39e-65 |
| SLC40A1 | DC | 0.571 | 5.45e-62 |
| STEAP3 | Fibroblast | 0.558 | 1.05e-58 |
| HFE | Fibroblast | 0.556 | 3.10e-58 |
| SLC40A1 | CD8_Tcell | 0.550 | 7.50e-57 |
| SLC40A1 | Macrophage | 0.542 | 9.71e-55 |
